# A genomic epidemiology study of multidrug-resistant *Escherichia coli, Klebsiella pneumoniae* and *Acinetobacter baumannii* in two intensive care units in Hanoi, Vietnam

**DOI:** 10.1101/2020.12.09.20246397

**Authors:** Leah W. Roberts, Le Thi Hoi, Fahad A. Khokhar, Nguyen Thi Hoa, Tran Van Giang, Cuong Bui, Tran Hai Ninh, Dao Xuan Co, Nguyen Gia Binh, Hoang Bao Long, Dang Thi Huong, James E. Bryan, Archie Herrick, Theresa Feltwell, Behzad Nadjm, H. Rogier van Doorn, Julian Parkhill, Nguyen Vu Trung, Nguyen Van Kinh, Zamin Iqbal, M. Estée Török

## Abstract

**Background:** Vietnam has high rates of antimicrobial resistance (AMR) but limited capacity for genomic surveillance. This study used whole genome sequencing (WGS) to examine the prevalence and transmission of three key AMR pathogens in two intensive care units in Hanoi, Vietnam.

**Methods:** A prospective surveillance study of all adults admitted to intensive care units (ICUs) at the National Hospital for Tropical Diseases (NHTD) and Bach Mai Hospital (BMH) was conducted between June 2017 and January 2018. Clinical and environmental samples were cultured on selective media, characterised using MALDI TOF MS, and illumina sequenced. Phylogenies based on the *de novo* assemblies (SPAdes) were constructed using Mafft (PARsnp), Gubbins and RAxML. Resistance genes were detected using Abricate against the NCBI database.

**Findings:** 3,153 *Escherichia coli, Klebsiella pneumoniae* and *Acinetobacter baumannii* isolates from 369 patients were analysed. Phylogenetic analysis revealed predominant lineages within *A. baumannii* (global clone [GC]2, sequence types [ST]2, ST571) and *K. pneumoniae* (ST15, ST16, ST656, ST11, ST147) isolates. Colonisation was most common with *E. coli* (88.9%) followed by *K. pneumoniae* (62.4%). Of the *E. coli*, 91% carried a *bla*CTX-M variant, while 81% of *K. pneumoniae* isolates carried *bla*NDM (54%) and/or *bla*KPC (45%). Transmission analysis using single nucleotide polymorphisms (SNPs) identified 167 clusters involving 251 (68%) patients, in some cases involving patients from both ICUs. There were no significant differences between the lineages or AMR genes recovered between the two ICUs.

**Interpretation:** This study represents the largest prospective surveillance study of key AMR pathogens in Vietnamese ICUs. Clusters of closely related isolates in patients across both ICUs suggests recent transmission prior to ICU admission in other healthcare settings or in the community.

**Funding:** This work was funded by the Medical Research Council Newton Fund, United Kingdom; the Ministry of Science and Technology, Vietnam; and the Wellcome Trust, United Kingdom.

**Research in context:** *Evidence before this study:* Globally, antimicrobial resistance (AMR) is projected to cause 10 million deaths annually by 2050. Ninety percent of these deaths are expected to occur in low- and middle-income countries (LMICs), but attributing morbidity and mortality to AMR is difficult in the absence of comprehensive data. Whilst efforts have been made to improve AMR surveillance in these settings, this is often hampered by limited expertise, laboratory infrastructure and financial resources.

*Added value of this study:* This is the largest prospective surveillance study of three key AMR pathogens (*E. coli, K. pneumoniae* and *A. baumannii*) conducted in critical care settings in Vietnam. Sampling was restricted to patients who were colonised or infected with extended spectrum beta-lactamase (ESBL) producing and/or carbapenem-resistant organisms. Colonisation with more than one organism was very common, with multidrug-resistant (MDR) *E. coli* being predominant in stool samples. A small number of predominant lineages were identified for *K. pneumoniae* and *A. baumannii*, while the *E. coli* isolates were highly genetically diverse. A large number of genomic clusters were identified within the two ICUs, some of which spanned both ICUs. There were no significant differences between lineages or AMR genes between the two ICUs.

*Implications of all the available evidence:* This study found high rates of colonisation and infection with three key AMR pathogens in adults admitted to two Vietnamese ICUs. Whilst transmission was common within ICUs the finding of similar lineages and AMR genes in both ICUs suggests that dissemination of AMR occurs prior to ICU admission, either in referral hospitals or in community settings prior to hospital admission. Strategies to tackle AMR in Vietnam will need to account for this by extending surveillance more widely across hospital and community settings.

## Introduction

Low- and middle-income countries (LMICs) have reported widespread antimicrobial resistance (AMR) in healthcare, community and agricultural settings. In South-East Asia, dense human populations, intensive animal farming, unrestricted access to antibiotics and limited laboratory infrastructure have all contributed to the rapid expansion of AMR (*1, 2*).

Much of this burden arises from excessive use of antimicrobials in human and animal populations. In Vietnam, antimicrobial usage has been estimated to be two times higher in humans, and 1.5 times higher in animals, as compared to the European Union (*3*). Despite legal restrictions in Vietnam, antibiotics are often dispensed without prescriptions in the community (*4*). Broad-spectrum antibiotics are also commonly administered in healthcare settings to mitigate the effects of limited capacity for microbiological testing and infection control (*4, 5*). Detection of both resistant bacteria and antimicrobials have been recorded in the environment (*6, 7*), hospital waste (*8*) and food sources (*9, 10*).

Extensive AMR has led to increased pressure on hospitals and is particularly problematic in critical care settings. Whilst AMR surveillance based on phenotypic testing in Vietnam has improved in recent years, the infrastructure required for systematic genomic surveillance remains to be established. Genomic analysis is important to determine circulating lineages and elucidate potential transmission events, as other methods do not provide the same level of resolution (*11*). Over the past decade, studies have demonstrated the utility of WGS in characterising AMR, transmission routes, and dominant lineages (*12-14*). LMICs however remain relatively understudied, with few studies conducted in Vietnamese hospitals (*15-18*).

In order to address this knowledge gap, we conducted a prospective genomic surveillance study of key AMR pathogens in two hospitals in Vietnam. We targeted intensive care unit (ICU) patients as we hypothesised that these would be most likely to have been treated with antibiotics and to harbour AMR pathogens. Furthermore, we focussed our analysis on the three most commonly isolated species (*Escherichia coli, K. pneumoniae*, and *A. baumannii*) that were extended-spectrum beta-lactamase (ESBL) producers and/or carbapenem-resistant. An additional in-depth analysis focussed primarily on a subset of the *K. pneumoniae* isolates was also performed in a separate project (Pham et al., personal communication).

## Methods

### Study design, setting and participants

This prospective observational cohort study was conducted in two hospitals, the National Hospital for Tropical Diseases (NHTD) and Bach Mai Hospital (BMH) in Hanoi, Vietnam, between June 2017 to January 2018. All adult patients admitted to the ICUs of the two hospitals were eligible for inclusion in the study. Screening specimens (stool/rectal swabs, urine, skin/wound swabs and sputum/tracheal aspirates) were collected from ICU patients on admission, on discharge and weekly during their ICU stay. Environmental samples were collected on a monthly basis.

### Laboratory methods

Specimens were cultured on selective media to identify extended-spectrum beta-lactamase producers and carbapenem-resistant organisms. Target organisms (*Escherichia coli, Acinetobacter baumannii* and *Klebsiella pneumoniae*) were identified using MALDI-TOF MS and stored at -80 °C prior to be being shipped to the University of Cambridge, United Kingdom, where they underwent antimicrobial susceptibility testing and DNA extraction.

### Sequencing and bioinformatic analysis

DNA extracts were transferred to the Wellcome Sanger Institute for library preparation and sequencing. Further details of laboratory methods including read quality control, genome assembly, phylogenetic analysis, antibiotic resistance gene detection, multilocus sequence typing (MLST) and transmission cluster analysis are available in the Supplementary Materials.

## Results

### Samples included in the study

Between June 2017 to January 2018, a total of 3,367 isolates were cultured, comprising *Escherichia coli* (n=765), *Klebsiella pneumoniae* (n=1,372) and *Acinetobacter baumannii* (n=1,230). Thirty-one isolates were excluded from the analysis because of poor assembly quality. A further 150 isolates were excluded because of suspected inter-species contamination, and 33 isolates were excluded because of suspected intra-species (strain-level) contamination (Supplementary Figure 2). Thus 3,153 isolates (93.6%), comprising 2,901 isolates from 369 patients, and 252 environmental isolates, passed quality filtering and were included in the final analyses.

### Clinical characteristics

3,153 bacterial isolates and 369 patients were included in the study. The number and type of samples and the participant baseline characteristics and outcomes are summarised in Table 1 and Supplementary Figure 3. The number of samples collected was higher, and the median length of ICU stay were longer at NHTD than BMH.

**Table 1:** Summary of study samples and participants.

|  | Bach Mai Hospital | National Hospital of Tropical Diseases |
| --- | --- | --- |
| **Samples** |  |  |
| Total | 1042 | 2111 |
| Clinical | 993 (95%) | 1898 (90%) |
| Environmental | 49 (5%) | 213 (10%) |
| **Study participants** |  |  |
| Number | 182 | 187 |
| Male sex | 104 (57%) | 114 (61%) |
| Female sex | 71 (39%) | 50 (27%) |
| Sex not recorded | 7 (4%) | 23 (12%) |
| **Age** |  |  |
| Male | 53 years (range 16-85) | 55.2 years (range 5-92) |
| Female | 53.4 years (range 19-91) | 55.7 years (range 10-90) |
| Age not recorded | n=5 (3%) | n=19 (10%) |
| **Duration of ICU admission** |  |  |
| Median length of stay (LOS) | 7.5 days (range 0 – 35) | 21 days (range 1 – 75) |
| LOS not recorded | n=4 (2.2%) | n=7 (3.7%) |
| **Outcome at ICU discharge** |  |  |
| Stable, discharged home | 10 (6%) | 38 (20%) |
| Improved, transferred to another ward | 117 (64%) | 83 (44%) |
| Deteriorated, transferred to another ward | 3 (2%) | 7 (4%) |
| Discharged home to die | 44 (24%) | 43 (23%) |
| Died in hospital | 4 (2%) | 9 (5%) |
| Not recorded | 4 (2%) | 7 (4%) |

All of the 369 patients were colonised or infected with one or more of the three species (*E. coli, A. baumannii* and *K. pneumoniae*): 146 patients (40%, 55 at BMH and 91 at NHTD) with all three species; 133 patients (36%, 66 at BMH and 67 at NHTD) with two of the three species; and 90 patients (24%, 61 at BMH and 29 at NHTD) only had one species detected.

Both *E. coli* and *K. pneumoniae* were isolated primarily from stool/rectal swabs (627/721 [87.0%] and 822/1316 [62.5%], respectively). *K. pneumoniae* was also isolated from other sites including sputum (325/1316 [24.7%]), urine samples (63/1316 [4.8%]) and skin swabs (17/1316 [1.3%]). In contrast, *A. baumannii* isolates were predominantly isolated from sputum (621/1116 [55.6%]), followed by stool/rectal swabs (247/1116 [22.1%]), urine (49/1116 [4.4%]) and skin swabs (36/1116 [3.2%]). *A. baumannii* also accounted for the highest number of environmental isolates (161/1116, [14.4%]), compared to 6.5% (85/1316) for *K. pneumoniae* and 2.2% (16/721) for *E. coli*.

### Whole genome sequencing reveals dominant circulating lineages

Phylogenetic trees for each species were constructed to explore lineage diversity. The *E. coli* isolates were found to be highly diverse, with isolates spread over eight phylogroups and 80 sequence types (STs) (Figure 1). The most prevalent ST was ST648 (phylogroup A; 11.8%), followed by ST410 (phylogroup C; 9.7%), ST617 (phylogroup A; 9.2%), ST131 (phylogroup B2; 7.9%) and ST1193 (phylogroup B2; 7.4%). Overall, 33 of the 80 STs only had one representative isolate in this dataset.

**Figure 1:**
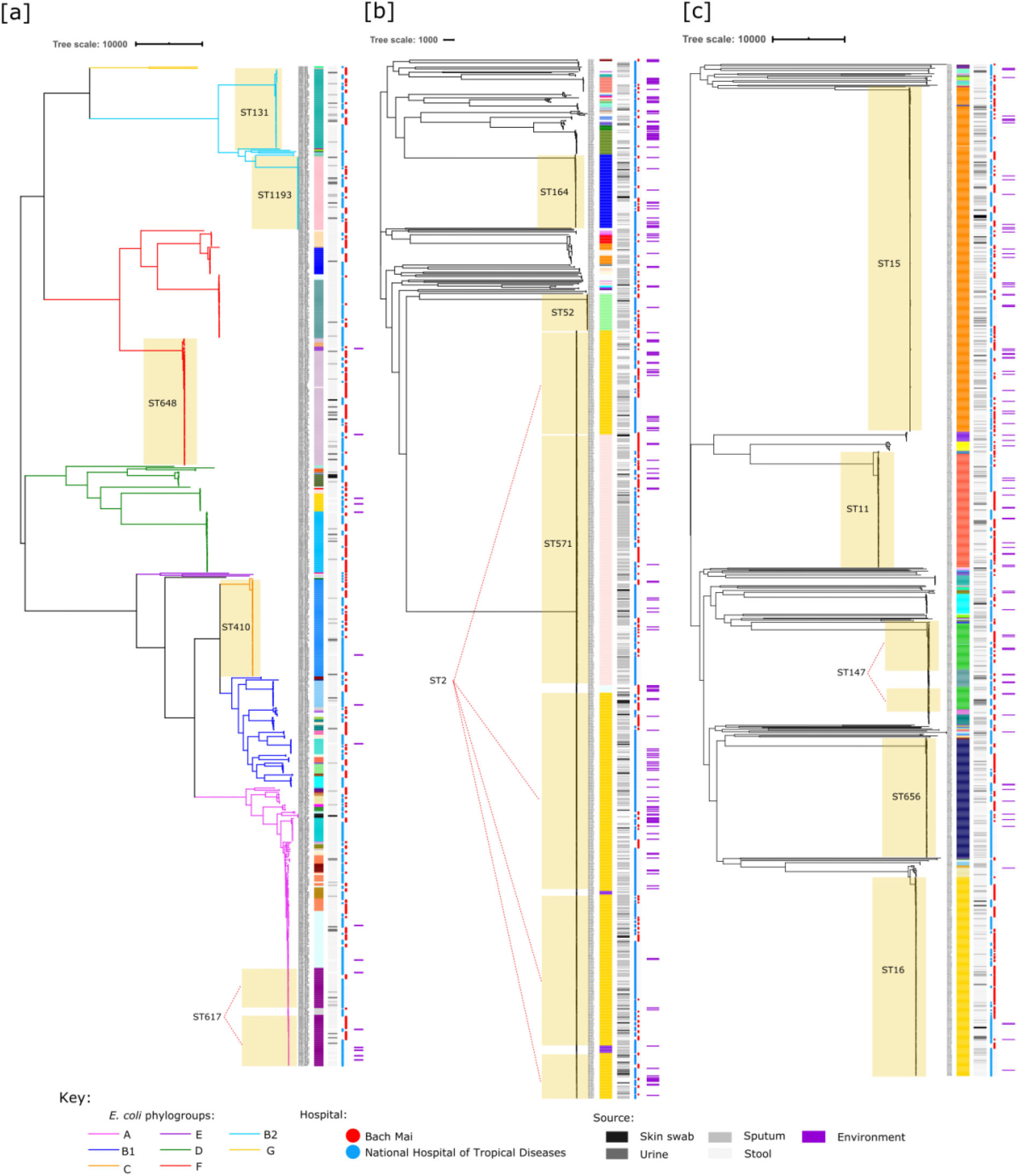
Whole genome phylogenies for [A] *E. coli*, [B] *A. baumannii*, and [C] *K. pneumoniae:* recombination-filtered core-SNP trees with mid-point root. Tree metadata includes (from left to right column beside trees): MLST, source and hospital. Outermost purple bars indicate environmental isolates. Branches corresponding to *E. coli* phylogroups are coloured accordingly. Main STs are highlighted in the image using the pale-yellow boxes.

In contrast, the *K. pneumoniae* and *A. baumannii* isolates appeared to have a limited number of dominant lineages. More than 80% of the *K. pneumoniae* isolates were from one of five STs, including ST15 (n=34%), ST16 (n=20%), ST656 (n=12%), ST11 (n=11%) and ST147 (n=7%). The majority of *A*. *baumannii* were global clone 2 (GC2) (n=832, 74.6%) (*19*) and mainly belonged to ST2 (n=48%) and ST571 (n=24%) (based on the Pasteur scheme). There did not appear to be any specific relationship between STs and hospitals, with all of the major ST lineages represented in both ICUs.

In order to gain broader insight into the lineages, we selected globally representative strains to contextualise our dataset. Addition of these global representatives into the *E. coli* phylogeny showed that most isolates belonged to a globally diverse set of STs that were not unique to Vietnam, but found across parts of North America, Europe and Asia (Supplementary Figure 4). Similarly, several of the major *K. pneumoniae* lineages were represented globally, particularly ST147, ST11 (mainly from China and the USA) and ST15 (mainly other Asian countries) (Supplementary Figure 5). However, it was also clear that local expansion was prominent, particularly among the ST656, ST16 and ST15 lineages. For *A. baumannii*, we focused primarily on GC2 isolates (Supplementary Figure 6). There was very little representation of global strains within our dataset, and those that were available consisted mainly of strains from other parts of Asia.

Closer inspection of the global representatives found several strains in each species that were closely related (<5 core SNPs) to isolates in our dataset (Supplementary Table 1). Most of these global representatives were also isolated in Asian countries, with agriculture implicated as a source, based on two closely related *E. coli* representatives originally isolated from poultry (biosample SAMEA104188722) and a farm worker in Vietnam (biosample SAMEA5277968). There were only two closely related representatives isolated outside Asia, from the United Kingdom and Australia (*E. coli* ST1193 and ST131, respectively, which are both well-characterised global lineages).

### High prevalence of antibiotic resistance genes among majority of isolates

Almost all isolates carried acquired resistance genes belonging to at least three antibiotic classes, with 90% of *E. coli*, 97% of *K. pneumoniae* and 41% *A. baumannii* carrying genes across five antibiotic classes (Figure 2). There were no discernible differences based on sample source or hospital, with the exception of *E. coli* detected in wound/skin swabs (n=6) which appeared on average to carry resistance to more antibiotic classes. *K. pneumoniae* isolates tended to fall into one of three “peaks” (Figure 2). This was due to lineage-specific carriage of acquired resistance genes, where ST15 isolates tended to carry resistance to more classes, compared to ST16 which often carried the least. The other three main lineages (ST656, ST11, ST147) fell between these two peaks. The exception was in the environmental samples, where only two peaks were seen and was likely due to very few ST16 isolates being detected in the environment.

**Figure 2:**
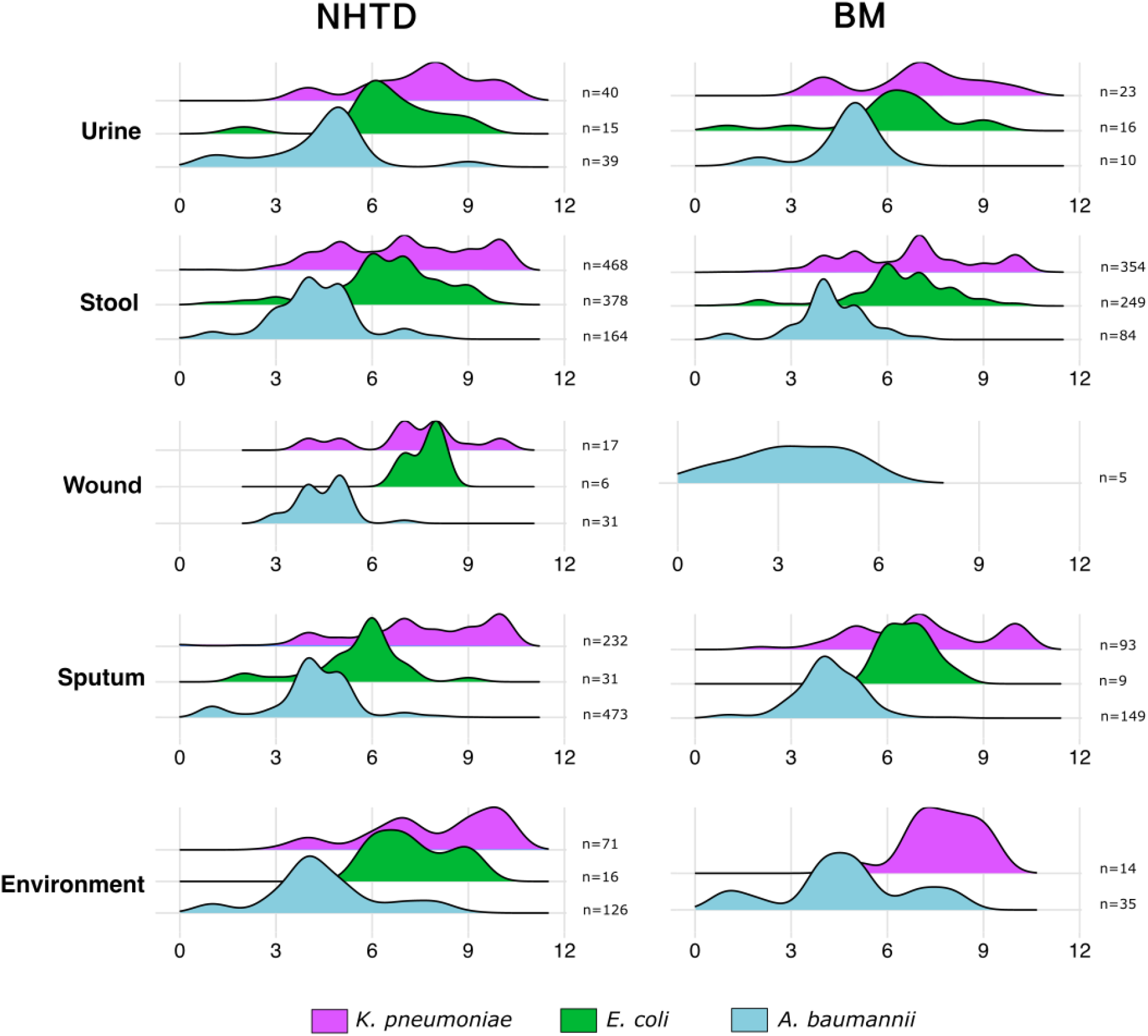
Summary of isolates and the number of antibiotic resistance classes separated by species, hospital and sample type.

Resistance to antibiotics classes varied across the *E. coli* phylogeny, reflective of the diversity of strains within the dataset (Supplementary Figure 7). *bla*CTX-M genes were found in most *E. coli*, with *bla*CTX-M-15 (36%), *bla*CTX-M-27 (30%) and *bla*CTX-M-55 (17%) the most prevalent (Table 2). *bla*KPC-2 (13%) and *bla*NDM-[1,4,5,7] (24%) were present sporadically across the phylogroups, suggesting independent acquisitions events. Only 4% (n=28) of isolates carried *mcr* genes conferring resistance to colistin. Again, these seemed to be independent acquisitions, with the exception of an ST206 cluster (phylogroup A; n=11) involving three patients from NHTD.

**Table 2.** Summary of resistance genes found in the three species.

| Resistance gene class | *E. coli* | *A. baumannii* | *K. pneumoniae* |
| --- | --- | --- | --- |
| Tetracycline | 563 (78.1%) | 701 (62.8%) | 753 (57.2%) |
| Sulphonamide | 649 (90%) | 746 (66.8%) | 969 (73.6%) |
| Fluoroquinolone | 161 (22.3%) | 19 (1.7%) | 1187<sup>3</sup> (90.2%) |
| Colistin | 28 (3.9%) | 0 (0%) | 10 (0.8%) |
| Fosfomycin | 48 (6.7%) | 5 (0.4%) | 1316<sup>3</sup> (100%) |
| MLS | 579 (80%) | 816 (73.1%) | 623 (47.3%) |
| Trimethoprim | 621 (86.1%) | 41 (3.7%) | 982 (74.6%) |
| Phenicols | 274 (38%) | 176 (15.8%) | 675 (51.3%) |
| Bleomycin | 179 (24.8%) | 36 (3.2%) | 717 (54.5%) |
| β-lactamase | 718 (99.6%) | 1018 (91.2%) | 1286 (97.7%) |
| *Class C:* |  |  |  |
| EC | 721<sup>1</sup> | 2 | 0 |
| ACT | 0 | 1 | 0 |
| CMY | 209 | 1 | 2 |
| DHA | 26 | 2 | 24 |
| *Class A:* |  |  |  |
| LAP | 17 | 0 | 142 |
| CARB | 0 | 63 | 0 |
| PER | 0 | 68 | 0 |
| TEM | 347 | 697 | 686 |
| SHV | 7 | 9 | 1292<sup>3</sup> |
| VEB | 0 | 12 | 3 |
| CTX | 613 | 4 | 681 |
| KPC | 94 | 3 | 593 |
| *Class D:* |  |  |  |
| OXA | 251 | 996<sup>2</sup> | 611 |
| *Class B:* |  |  |  |
| IMP | 0 | 6 | 1 |
| NDM | 173 | 35 | 716 |
| Rifamycin | 114 (15.8%) | 63 (5.6%) | 810 (61.6%) |
| Aminoglycoside | 680 (94.3%) | 1115 (99.9%) | 1290 (98%) |
| Streptothricin | 11 (1.53%) | 8 (0.7%) | 0 (0%) |

<sup>1</sup>blaEC intrinsic in *E. coli*
<sup>2</sup>blaADC and blaOXA intrinsic in *A. baumannii* (except OXA-[1,10,23,58,72])
<sup>3</sup>fosA (fosfomycin), oqxAB (fluoroquinolone) and blaSHV intrinsic in *K. pneumoniae*
ESBL resistance genes are highlighted in grey and carbapenem resistance genes in yellow

Conversely, MDR gene presence across the *K. pneumoniae* isolates appeared consistent with the main lineages, suggesting clonal expansion rather than diverse sampling of the species (Supplementary figure 8). Similar to the *E. coli*, incidence of *bla*CTX-M-15 (37.5%) was high, but less so than *bla*KPC-2 (45%) and *bla*NDM (54%) (NDM-4 [27.9%], NDM-1 [24.7%] and NDM-5 [1.8%]) (Table 2).

Acquired AMR genes were overall less prevalent among the *A. baumannii* isolates. Similar to the *K. pneumoniae*, resistance to specific classes tended to be a feature of each distinct lineage, suggesting clonal expansion (Supplementary Figure 9). The carbapenemase gene *bla*OXA-23 was present in 83% of the dataset, with *bla*OXA-58 and *bla*OXA-72 present at much lower frequencies (5% and 0.2% respectively) (Table 2). The aminoglycoside resistance gene *armA* was also highly prevalent (76%).

Overall, 133 AMR genes were detected in BM and 154 were detected in NHTD. 49 genes were unique to either hospital (35 in NHTD, 14 in BM), but were only detected at a prevalence of less than 0.1%, suggesting sporadic cases. The remaining 129 genes were the same across both hospitals. Genes with at least 1% prevalence in each hospital were found to be almost identical. There were only five exceptions where the gene prevalence was >1% in BM but <1% in NHTD (*bla*NDM-4 (0.98%), *dfrA12* (0.92%), *rmtB1* (0.86%), *qnrB6* (0.74%) and *bla*OXA-181 (0.56%)). We also attempted to discover possible mobile genetic element (MGE)-driven AMR transmission by monitoring fluctuation of AMR genes prevalence over time (further explained in the supplementary results, supplementary figure 10).

### Most patients carried several AMR strains

For patients already infected or colonised with *E. coli, K. pneumoniae* and/or *A. baumannii*, over half had repeat isolation of the same bacterial species at a later timepoint during their stay (*E. coli*: 138/270 [51%], *K. pneumoniae*: 169/294 [57%], *A. baumannii*: 133/230 [58%]). Of these patients, 60 to 70% had different sequence types (ST) (*E. coli*: 67%, *K. pneumoniae*: 68%, *A. baumannii*: 64%) (Figure 3). For *E. coli* and *K. pneumoniae*, the majority of patients only had isolates detected in stool (80% and 54%, respectively). Conversely, most patients with *A. baumannii* were detected only in sputum (38%), or sputum and stool (32%).

**Figure 3:**
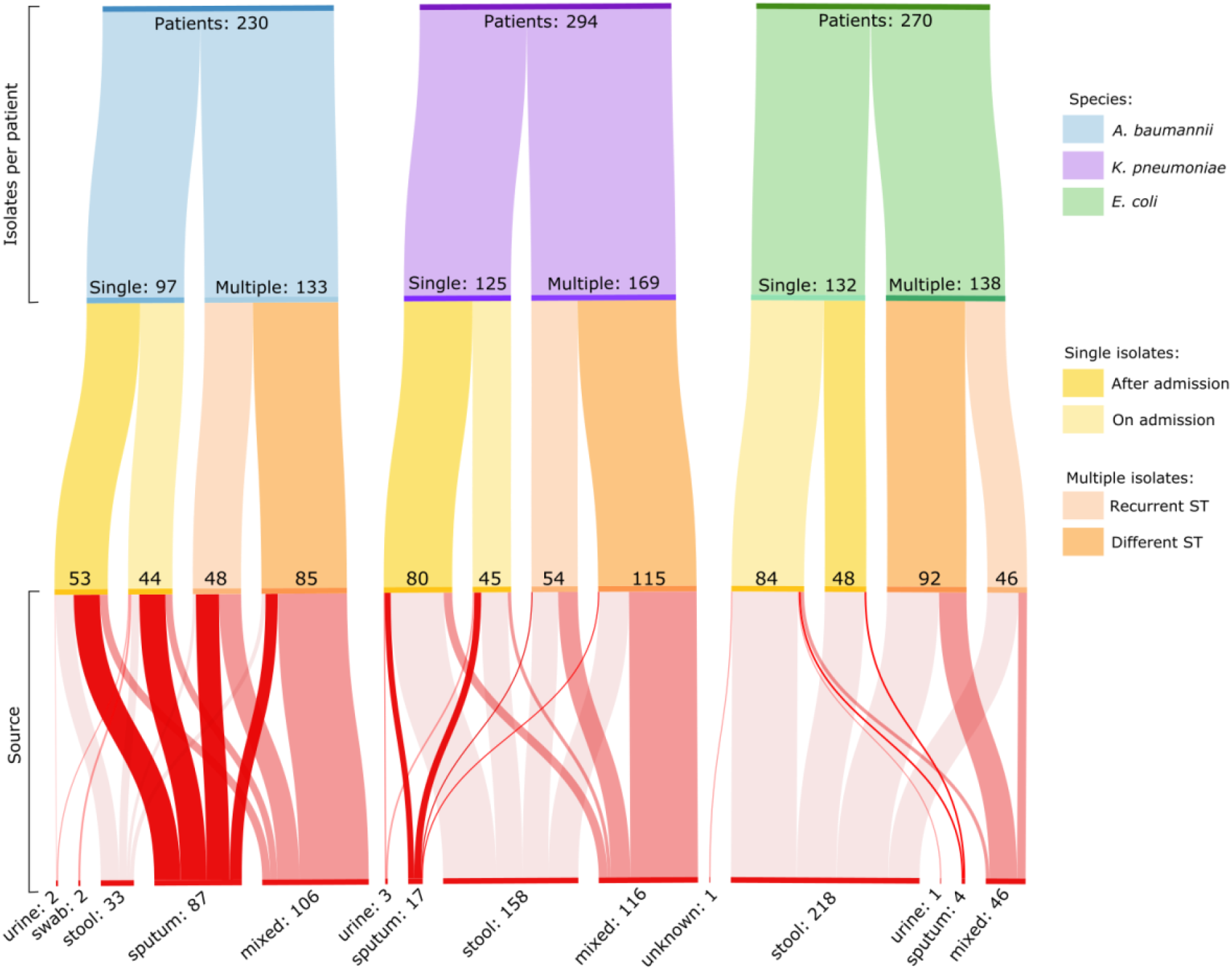
Overview of strain diversity, recurrence and source among study patients: “Patients” refers to the total number of patients in this study that had at least one isolate of that species. Within each species, we evaluated whether patients had (i) only a single isolate for that species, or (ii) multiple isolates. If only a single isolate, we determined whether it was collected on admission to the ICU or after. For multiple isolates, we determined if the patient’s isolates were the same ST (recurrent) or a different ST. We finally looked at how many patients had isolates from one site (urine, swab, stool or sputum) or mixed sites (any combination of sites).

### Evidence of extensive transmission between ICU patients

Temporal observation of the isolates found no obvious association of any time point with any ST to suggest an outbreak of a specific lineage. In order to investigate potential clusters within the ICUs at a higher resolution, we examined plausible short-term transmission events using single nucleotide polymorphisms (SNPs).

To identify closely related strains that could indicate recent transmission, we evaluated clusters based on SNP distances across the core genome of each species for this dataset. Given the short sampling period, none of the three major species were likely to acquire more than 1 SNP while in the hospital. As such, we looked at samples with genomic evidence of most recent transmission; zero SNP clusters (follow-up with higher thresholds detailed below). Clusters were defined when they involved >1 patient. Clusters involving a single patient and environmental samples were not included.

Most clusters were detected in *K. pneumoniae* and *A. baumannii* isolates, with 71 and 74 clusters representing 38% and 52% of total isolates for that species, respectively (Figure 4). *K. pneumoniae* had some of the largest clusters, ranging in size from 2 to 79 isolates, while *A. baumannii* clusters were smaller, between 2 to 33 isolates. Only 22 clusters were detected in *E. coli* and were generally small (median 3 isolates, range 2 to 9 isolates), representing only 13% of the *E. coli* dataset.

**Figure 4:**
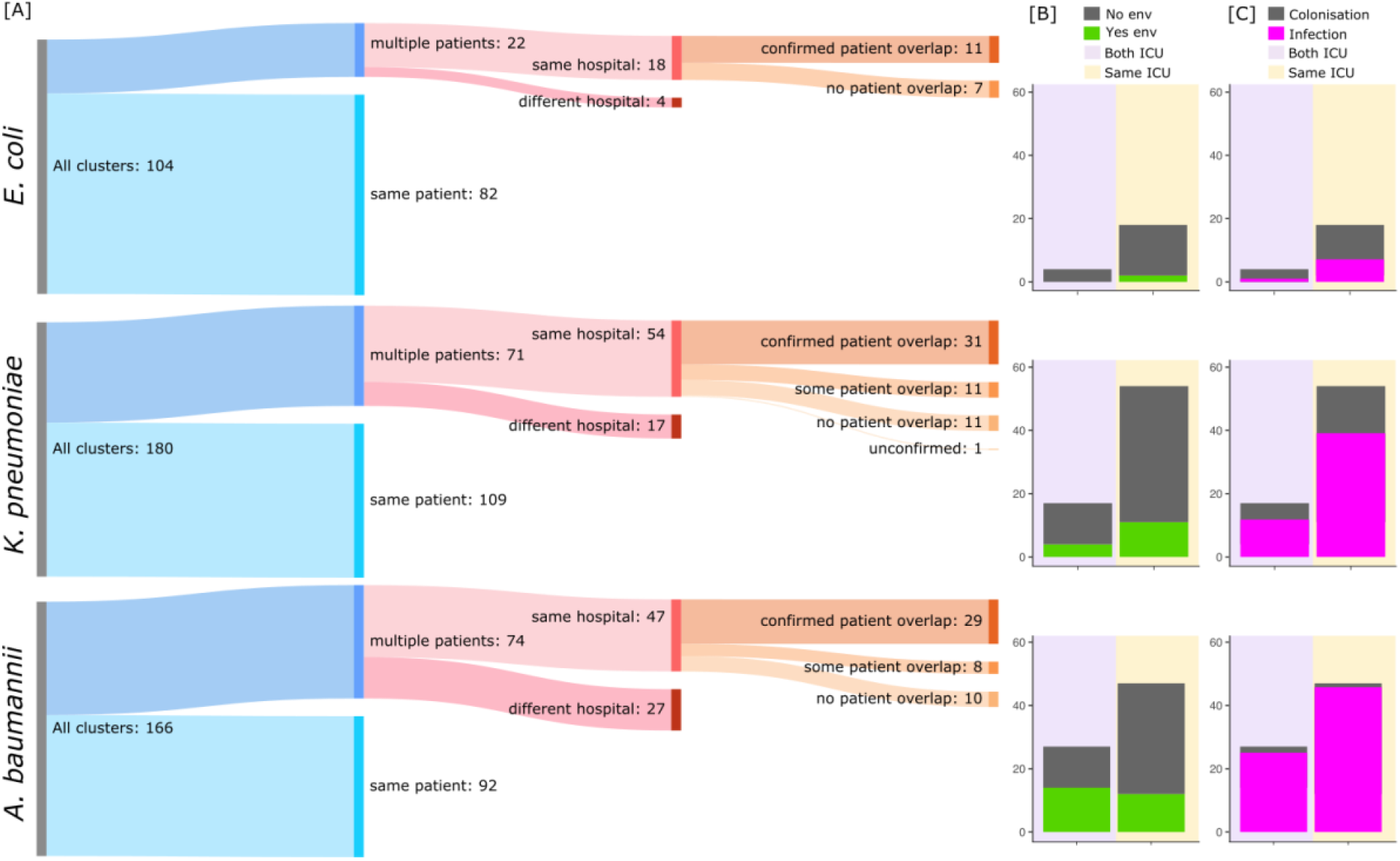
Summary of 0 SNP clusters in all species: **[A]** Clusters were defined as (i) multiple patients: samples were derived from at least two different patients, or (ii) same patient: isolates were derived from the same patient, or only a single patient and the environment. Epidemiological evidence to support clusters was defined as (i) confirmed patient overlap: all patient ICU stays overlap with another in the same cluster, (ii) some patient overlap: at least 2 patient ICU stays overlap, and (iii) zero patient overlap between all patients in cluster. **[B]** Environmental isolates in clusters: clusters were counted if an environmental isolate was found in that cluster. Y-axis = number of clusters. **[C]** Colonisation vs. infection: clusters were counted if they (a) had only isolates from stool (i.e. colonisation) or (b) had isolates from urine, swabs and/or sputum with or without isolates from stool (i.e. infection). Y-axis = number of clusters.

For all three species, the majority of clusters were detected between patients within a single ICU. Evaluating admission and discharge dates further confirmed patient overlap in these clusters (Figure 4, Supplementary Figures 12 to 17). Additionally, between-ICU clusters were also detected (discussed further below).

Compared to *E. coli* and *K. pneumoniae, A. baumannii* clusters were more often associated with environmental isolates (24/74 clusters, Figure 4). The largest *A. baumannii* cluster within the same hospital ICU involved 33 isolates from eight patients and eight environmental samples (Supplementary Figure 13; ST451, cluster number 13). Admissions for patients in this cluster overlapped with detection of the same strain in the environment, which was also detected in the hospital environment several months later. Only two *E. coli* clusters contained related environmental isolates. *K. pneumoniae* environmental isolates were more often found in within-hospital (ICU) clusters (n=11) compared to between-hospital clusters (n=4).

To broadly evaluate infection risk for each outbreak cluster, we determine whether clusters contained (i) stool sample / rectal isolates only, indicating colonisation or (ii) isolates from urine, skin swab or sputum samples, which could represent infection or colonisation at multiple sites. The majority of *A. baumannii* clusters contained isolates from non-stool / rectal swab samples (Figure 4). Conversely, colonisation-only clusters were common for *E. coli*; the largest *E. coli* cluster involving both hospital ICUs contained only 5 isolates from four patients, which were all isolated from stool (Supplementary Figure 14; ST617, cluster number 37). *K. pneumoniae* clusters were a mix, with both colonisation-only clusters and infection clusters.

In addition to suspected within-ICU transmission, we also detected a number of clusters involving patients from both hospital ICUs (Figure 4). The most pronounced example of this was a large ST15 *K. pneumoniae* cluster involving 79 isolates from 38 patients and 6 environmental samples (Supplementary Figure 18). Most were collected between July to September, with some late occurrences in October and November. All patients from NHTD between July to September had overlapping timelines, consistent with spread within the ICU (Supplementary figure 16; ST15, cluster number 15). Only one patient had no evidence of overlap (ND162) but did cluster with environmental isolates from the same timeframe, indicating a possible environmental source. Patients from BM appeared to have both patient overlap and consecutive acquisitions without patient overlap as time progressed. This suggests that patient-to-patient transmission and also transmission via other routes (e.g. inadequate cleaning before the next patient, transmission via healthcare workers) may have been important factors in the spread of this strain.

The identification of closely related isolates between independently operating ICUs suggested that there may have been a common source located outside the ICU e.g admission to the same location prior to admission to ICU. To determine whether certain lineages were more associated with acquisition within the ICU, we assessed diversity on arrival (i.e. the patients’ first sample) versus diversity within the ICU (all other samples). Based on ST alone, we found a slight increase in diversity in the ICU versus on arrival (Supplementary Figure 19). However, the unique STs recovered in either setting only represented a small portion of the isolates overall. All of the main lineages for each species were found on both admission and within the ICU (Supplementary Figure 19 and 20).

### Identification of multiple transmission clusters per patients

Overall, there were 251 patients (representing 68% of the cohort) involved in 167 clusters across the three species collected over the course of this study. 112 patients were only involved in a single cluster during their time in the ICU (Figure 5). However, the remaining 139 patients were involved in at least two clusters, with one patient involved in 12 clusters. For patients with at least two clusters, 20 had clusters from all three species, 94 had clusters from two species and 25 had only one species. Overall, we saw a general trend towards more clusters in a single patient as they spent more time in the ICU ward.

**Figure 5:**
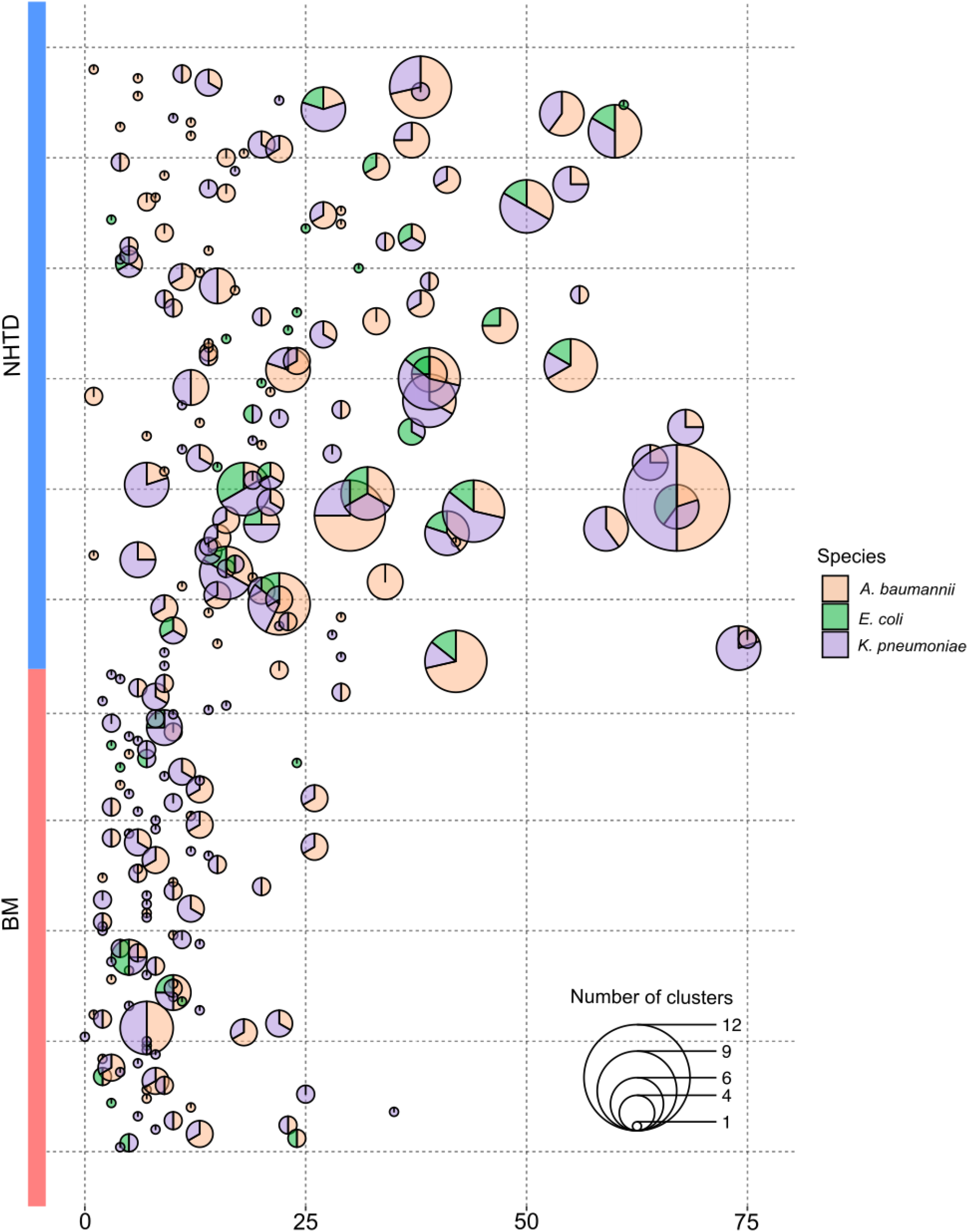
Scatterpie showing the number of clusters in patients across all species: y-axis represents patients from BM or NHTD involved in at least one 0 SNP cluster. X-axis represents length of stay for that patient; one pie is plotted per patient at the duration of their stay. Each circle represents 0 SNP clusters in a single patient. The size of the clusters corresponds to the number of clusters, while the colour corresponds to the species.

To determine if any of our (zero SNP) clusters were potentially derived from a single original cluster predating their time in the ICU, we looked at SNP distances between clusters of the same ST (Supplementary Figure 21). At a threshold of five SNPs, several of the prominent STs within each species formed large clusters, including ST804 in *A. baumannii* and ST16 in *K. pneumoniae*. At this threshold, we found 29 clusters in the *A. baumannii* dataset (originally 74), 23 clusters in the *K. pneumoniae* (originally 71) and 19 clusters in *E. coli* (originally 22). By readjusting our analysis per patient using these clusters, we found that slightly more patients (123 versus 112 previously) may have only been involved in a single cluster during their stay (Supplementary figure 22). 128 patients had 2 clusters, with the maximum number of clusters in a single patient being 7 (n=3 patients).

## Discussion

Here we present a large prospective genomic surveillance study of ESBL- or carbapenemase-producing organisms from three key AMR pathogens from two hospital ICUs in Vietnam. We used WGS to capture a high-resolution snapshot of the dominant circulating lineages over a six-month period. In this study we focused on *E. coli, K. pneumoniae*, and *A. baumannii*, as these were the most commonly isolated species from both ICUs. These three species have also been reported as highly prevalent in other Vietnamese studies (*20*), and are amongst the most clinically significant Gram-negative bacteria, having been designated as “critical” priority pathogens for research and development of new antibiotics by the World Health Organisation (*21*).

Despite limiting our study samples to ESBL-producing and/or carbapenem-resistant isolates belonging to three species, we identified a large number of isolates (an average of 17 and 10 isolates/day from patients in BMH and NHTD respectively). This is exceedingly high compared to other countries, such as the United Kingdom (UK), where a similar study only found 199 ESBL-producing Enterobacteriaceae over the course of one year (0.5 isolates/day) from three hospital sites between 2008 to 2009 (*22*). A point-prevalence survey conducted in a UK hospital in 2017 also identified no positive carbapenemase-producing Enterobacteriaceae (CPE) from 540 samples (*23*).

Colonisation was found to be a large reservoir for AMR, with the majority of *E. coli* and more than half of the *K. pneumoniae* isolated from stool samples, as has been documented previously in Vietnamese hospitals (*24*). High community usage of antibiotics in Vietnam is likely to promote colonisation with AMR bacteria, as prior treatment with antibiotics is known to lead to colonisation (*25*). Colonisation itself has been identified as a risk factor for subsequent infection (*26-28*), and has been previously documented in ICU patients in southern Vietnam (*29*). Colonisation with AMR *E. coli* has also been identified as a risk for transferral of resistance to other colonising pathogens, such as *Shigella* (*30*).

In addition to evidence for recent transmission between patients on the same ICU, we also identified clusters involving patients from both hospital ICUs. This result was unexpected, as there was no direct transfer of patients between the two ICUs. The most likely explanation for these clusters is a source outside of the ICUs. This might be other wards or hospitals which may have referred patients to intensive care, or AMR strains that have been acquired in the community, reflecting high rates of antibiotic use. Based on the similarity between lineages and AMR across both ICUs, we suggest that transmission, particularly of the predominant *A. baumannii* and *K. pneumoniae* lineages, is likely already circulating outside of the ICUs, where it is then further propagated.

In this context, we should potentially reconsider how we model AMR surveillance and control. Knowing whether hospitals are the primary source of AMR bacteria (and subsequent transmission into the community) (*31*), or whether the high rates are part of a more general, endemic pattern of circulating resistant strains is important for our understanding of local transmission dynamics. If the latter is true, this leads to very different national and indeed global management plans.

We acknowledge some limitations to our study. Firstly, we did not extensively explore plasmid profiles amongst the samples because of the limitations of short read sequence data. Secondly, although serial samples were collected and cultured, we selected single colony picks for sequencing. It is therefore possible that some diversity may have been lost at different timepoints during the study sampling period. Finally, this study focused on patient and environmental samples only. We were therefore unable to investigate potential transmission events involving hospital staff and/or visitors.

Nevertheless, we present the largest prospective surveillance study of multidrug-resistant *E. coli, A. baumannii* and *K. pneumoniae* in Vietnamese critical care patients to date. The extensive transmission and AMR detected within and between ICU wards suggests dominant circulating lineages of *A. baumannii* and *K. pneumoniae* existing both within hospitals, and potentially in community settings in Vietnam. Further work is required to expand genomic surveillance in hospital and community settings in order to inform AMR control strategies in Vietnam.

## Data Availability

Genome sequence data have been deposited in the European Nucleotide Archive (ENA) under the Bioproject PRJEB29424. A list of the sample accession numbers is available in Supplementary Dataset 1. Isolate genome assemblies (heterogenous sites masked and unmasked) are available on Figshare under the following DOI: 10.6084/m9.figshare.13303253,
10.6084/m9.figshare.13302728.

https://www.ebi.ac.uk/ena/browser/text-search?query=PRJEB29424

## Data sharing

Genome sequence data have been deposited in the European Nucleotide Archive (ENA) under the Bioproject PRJEB29424. A list of the sample accession numbers is available in Supplementary Dataset 1. Isolate genome assemblies (heterogenous sites masked and unmasked) are available on Figshare under the following DOI: 10.6084/m9.figshare.13303253, 10.6084/m9.figshare.13302728.

## Ethical statement

The study protocol was approved by the Scientific and Ethical Committees of the National Hospital for Tropical Diseases and Bach Mai Hospital and by the University of Cambridge Human Biology and Research Ethics Committee (reference: HBREC 2017.09). Written informed consent was obtained from the patient or from their relative prior to enrolment in the study.

## Author contributions

Conceptualisation: LWR, JP, NVK, ZI, MET

Data collection: FK, NTHM, LTH, NGB, DXC, LTH, NTH, TG, CB, HNT, BN, RvD, NVT

Sample processing: NTH, FK, JB, JH, TF

Methodology: LWR, ZI

Formal Analysis: LWR, ZI

Writing (original draft): LWR

Writing (review/editing): ZI, MET

Supervision: NVT, RvD, NVK, ZI, MET

Project administration: LTH, NVT, RvD, MET

Funding acquisition: MET, NVK and JP

## Acknowledgements

The authors would like to thank the patients for participating in this study and the clinical and laboratory staff of the National Hospital for Tropical Diseases and Bach Mai Hospital for their assistance with this study. We also acknowledge the sequencing team at the Wellcome Sanger Institute for their assistance with sequencing the samples including in the study.

