## Supplementary materials for "A genomic epidemiology study of multidrug-resistant *Escherichia coli, Klebsiella pneumoniae* and *Acinetobacter baumannii* in two intensive care units in Hanoi, Vietnam"

### Supplementary Methods

*Study design, setting and participants*

This prospective observational cohort study was conducted in two hospitals, the National Hospital for Tropical Diseases (NHTD) and Bach Mai Hospital (BMH) in Hanoi, Vietnam, between June 2017 to January 2018. All patients (aged 18 years or older) admitted to the adult ICUs of the two hospitals were eligible for inclusion in the study. NHTD is a specialist hospital for infectious and tropical diseases with a 22-bedded ICU which receives up to 400 patients per year. BMH is a large tertiary referral hospital, with a 45-bedded ICU that receives up to 1,200 patients per year. Both hospitals are located in the same area of Hanoi but operate independently of each other and do not share laboratory facilities, equipment or staff. Patients are not commonly transferred between the two hospitals.

*Study procedures*

Screening specimens were collected from ICU patients on admission, on discharge and weekly during their ICU stay. Specimens included stool/rectal swabs, urine, skin/wound swabs and sputum/tracheal aspirates. Environmental samples were collected using flocked swabs (from door handles, bed rails, medical equipment and patient tables) on a monthly basis. Clinical data related to the ICU admission were collected from the medical records and entered into a case record form and then into an electronic database. Laboratory data were collected and recorded in an electronic database.

*Laboratory methods and sequencing*

All patient and environmental specimens were cultured on selective media (CHROMagar™ ESBL, CHROMagar™ mSuperCARBA™, CHROMagar™ VRE, CHROMagar, France). Single colony picks of target organisms (*Escherichia coli*, *Acinetobacter baumannii* and *Klebsiella pneumoniae*) were selected and identified using MALDI-TOF MS (Bruker Diagnostics, Bremen, Germany) and stored at -80 °C. Stored isolates were shipped in two batches to the University of Cambridge, United Kingdom, where they were sub-cultured, re-identified using MALDI-TOF MS, and underwent antimicrobial susceptibility testing (Vitek-2, BioMérieux, Marcy L'Étoile, France). Isolate DNA was extracted using QIACube and the QIAamp 96 DNA QIACube HT kit (Qiagen, Hilden, Germany) prior to shipping to the Wellcome Sanger Institute for sequencing. DNA was sequenced in two batches on an Illumina HiSeq X10 machine (Illumina Inc., San Diego (CA), USA).

*Read quality control*

Raw Illumina reads were checked for quality using fastQC (v0.11.8) (*1*) and MultiQC (v 1.0.dev0) (*2*). Raw Illumina reads were also checked for contamination using Kraken2 (v2.0.7-beta) (*3*) and Bracken (v2.5) (*4*).

*Assembly*

Illumina reads were *de novo* assembled using SPAdes (v3.13.1) (*5*) and checked for quality using Quast (v5.0.2) (*6*) and CheckM (v1.0.18) (*7*). Suspected contamination was removed from the assemblies by removing contigs which were both supported by less than 10x coverage and were less than 100 base pairs (bp) in length. Assemblies that retained more than 1000 contigs after this initial filtering stage were removed. Other assemblies that were >6 Mbps (or >7 Mbps for *Pseudomonas* species) or had >500 contigs were checked for contamination using CheckM (v1.0.18) (*7*). Any assemblies that were found to have >5% contamination based on CheckM were removed. Reads were then remapped to filtered assemblies to check for remaining heterogenous (het) sites. Isolates that had >0.9% het sites were removed from the study. A complete list of isolates is provided in Supplementary Dataset 1.

*Phylogenetic construction*

Prior to phylogenetic construction, reads were mapped to filtered assemblies and sites that had <90% consensus compared to the reference allele were masked. Masked assemblies were then aligned using PARsnp (v1.2) under default settings (*8*). Core multi-alignments produced using PARsnp were filtered for recombination using Gubbins (v.2.3.5) (*9*). Phylogenies were then constructed using RAxML (GTR-GAMMA model) (v8.2.12) (*10*) as implemented through Gubbins. *E. coli* phylogroups were determined using ClermonTyping (*11*). Trees were visualised using iTol v5.6.2 (*12*). Details on global reference selection are provided in the Supplementary Methods.

*Antibiotic resistance gene detection*

Resistance genes and plasmid replicons were detected from draft assemblies using Abricate (v1.0.1) (*13*) and the NCBI database (for resistance genes) (*14*)**.** Genes were considered present if there was 90% coverage at 90% nucleotide identity (*15*).

*Multi-locus sequence typing (MLST)*

Sequence types were determined using mlst (v2.19.0) (https://github.com/tseemann/mlst) and the associated species scheme (specifically the Pasteur scheme for *A. baumannii*) (*16-18*).

*Transmission cluster analysis*

Transmission clusters were constructed using Transcluster (using the makeSNPClusters method, which ignores time of sampling and uses a pure SNP-distance cut-off) (*19*) using single nucleotide polymorphisms (SNPs) determined after recombination filtering using Gubbins. Transmission cut-offs were evaluated based on intra- and inter-patient SNP diversity within each species phylogeny (Supplementary Figure 1).

*Global reference selection:*

Global references for each of the three species were selected as follows:

1. *A. baumannii*: All complete *A. baumannii* available on NCBI as of the 22[nd] June 2020 were downloaded. Raw reads from (*20*) were also downloaded. Raw reads were checked for quality using FastQC (v0.11.8) and multiQC (v1.6). Reads were trimmed with Trimmomatic (v0.39) (*21*) under the following settings: LEADING:20 TRAILING:20 SLIDINGWINDOW:4:15 MINLEN:100. Trimmed reads were then *de novo* assembled using Spades (v3.13.1). Only isolates within global clone (GC)2 were included. A total of 175 genomes were added.
2. *K. pneumoniae*: Assemblies available from (*22*) were downloaded. Additionally, all complete *K. pneumoniae* available on NCBI as of the 22[nd] June 2020 were downloaded. A total of 798 genomes were added.
3. *E. coli*: PopPUNK (v1.1.3) (*23*) was used to assign all *E. coli* in this dataset to clusters in a predefined global *E. coli* collection (*24*). For each isolate that clustered, we used Mash (v2.2) (*25*) to determine the closest reference within the global dataset, resulting in 94 unique reference strains. For every *E. coli* ST not represented by these 94, we selected an *E. coli* reference from Enterobase (v1.1.2) (*26*), with a preference for (i) Vietnamese/Asian origin and/or (ii) human samples. A total of 140 genomes were added.

A list of all global references used is provided in Supplementary Dataset 2.

*Capsule typing:*

K and O antigens for *K. pneumoniae* and *A. baumannii* isolates were determined *in silico* using Kaptive (v0.5.1) (*27-29*). Matches that had a confidence of “low” or “none” were marked on the corresponding figures.

*Kleborate*:

*K. pneumoniae de novo* assemblies were analysed using Kleborate (v0.4.0-beta) (*30*) to confirm species, determine virulence profiles and detect *mgrB* insertions.

*Multi-locus sequence typing (MLST) for* Acinetobacter baumannii*:*

MLST for *A. baumannii* was additionally carried out using mlst (v2.19.0) (https://github.com/tseemann/mlst) and the Oxford Scheme (*31*). This publication made use of the PubMLST website (https://pubmlst.org/) developed by Keith Jolley (*32*) and sited at the University of Oxford. The development of that website was funded by the Wellcome Trust.

**Supplementary Results**

*Determining the role of mobile genetic elements in spreading AMR genes within the ICU*

In order to determine if certain time points throughout the study had different gene burdens (potentially indicative of mobile genetic element [MGE]-mediated transmission), we plotted gene presence versus date for genes equivalent to 1% prevalence in either hospital (Supplementary Figures 10). Overall, we found that both ICUs had a consistently high burden, making it difficult to distinguish significant gene fluctuations over time. In NHTD, we observed three genes that seemed to peak between November to December 2017. Examination of our dataset for isolates with these three genes (*bla*NDM-4, *bla*OXA-181 and *rmtB1*) revealed a subset of ST16 *K. pneumoniae* that carried all three genes as well as a single ST11 isolate from BM. We plotted the presence of these isolates over time, which mirrored the rise in prevalence in NHTD over the November to December period (Supplementary Figure 11). No *E. coli* nor *A. baumannii* isolates in this dataset carried all three genes.

**Supplementary Tables**

**Supplementary Table 1: Global representative strains found to be closely related to study isolates**

| Reference strain | Study isolate | Country | Year | Citation |
|---|---|---|---|---|
| ***K. pneumoniae*** | | | | |
| 1690099 | 26975_4#120 | Laos | 2015 | Wyres 2020 GenomeMed |
| COMRU-KPN-BC-2017-3 | 26968_6#88 | Cambodia | 2017 | |
| 201213-39019 | 26968_6#148 | Vietnam | 2013 | |
| 18-2374 | 27097_7#3 | South Korea | 2018 | CP041927.1 |
| PIMB15ND2KP27 | 27097_7#3 | Vietnam | 2015 | CP041639.1 |
| WCHKP7E2 | 26975_5#22 | China | 2015 | CP028806.2 |
| ***A. baumannii*** | | | | |
| DRR035557 | 26994_7#31 | Vietnam | 2011 | Tada et al 2015 BMC Infect Dis |
| DRR035584 | 29050_1#131 | Vietnam | 2012 | |
| DRR035551 | 26975_4#24 | Vietnam | 2011 | |
| DRR035554 | 26975_4#24 | Vietnam | 2011 | |
| DRR035570 | 26975_4#24 | Vietnam | 2011 | |
| DRR035613 | 26975_4#24 | Vietnam | 2013 | |
| DRR035545 | 29050_1#131 | Vietnam | 2011 | |
| DRR035583 | 29050_1#131 | Vietnam | 2012 | |
| AC29 | 26968_5#156 | Malaysia | 2011 | CP007535.2 |
| AC30 | 26968_5#156 | Malaysia | 2011 | CP007577.1 |
| Aba | 26968_5#164 | China | 2016 | CP030083.1 |
| DRR035531 | 26975_4#24 | Vietnam | 2011 | Tada et al 2015 BMC Infect Dis |
| DRR035538 | 26975_4#24 | Vietnam | 2011 | |
| DRR035562 | 26975_4#24 | Vietnam | 2011 | |
| DRR035564 | 26975_4#24 | Vietnam | 2011 | |
| DRR035611 | 26975_4#24 | Vietnam | 2012 | |
| DRR035579 | 29050_1#131 | Vietnam | 2012 | |
| DRR035593 | 26968_5#150 | Vietnam | 2012 | |
| DRR035595 | 26968_5#150 | Vietnam | 2012 | |
| DRR035603 | 26968_5#150 | Vietnam | 2012 | |
| DRR035532 | 26975_4#24 | Vietnam | 2011 | |
| DRR035534 | 26975_4#24 | Vietnam | 2011 | |
| DRR035536 | 26975_4#24 | Vietnam | 2011 | |
| DRR035539 | 26975_4#24 | Vietnam | 2011 | |
| DRR035547 | 26975_4#24 | Vietnam | 2011 | |
| DRR035563 | 26975_4#24 | Vietnam | 2011 | |
| DRR035587 | 26975_4#24 | Vietnam | 2012 | |
| DRR035602 | 26975_4#24 | Vietnam | 2012 | |
| DRR035540 | 26975_5#132 | Vietnam | 2011 | |
| DRR035591 | 26994_6#22 | Vietnam | 2012 | |
| ***E. coli*** | | | | |
| SRR3578916 | 26994_7#100 | United Kingdom | 2016 | Horesh et al Biorxiv 2020 |
| SAMEA104188722/ PRJEB21997 | 29109_2#65 | Vietnam (poultry) | 2017 | |
| esc_ca2906aa_as | 29339_1#373 | Australia | 2010 | |
| SAMEA5277968/ PRJEB30991 | 27261_7#149 | Vietnam (farmer) | 2015 | |

### Supplementary Figures

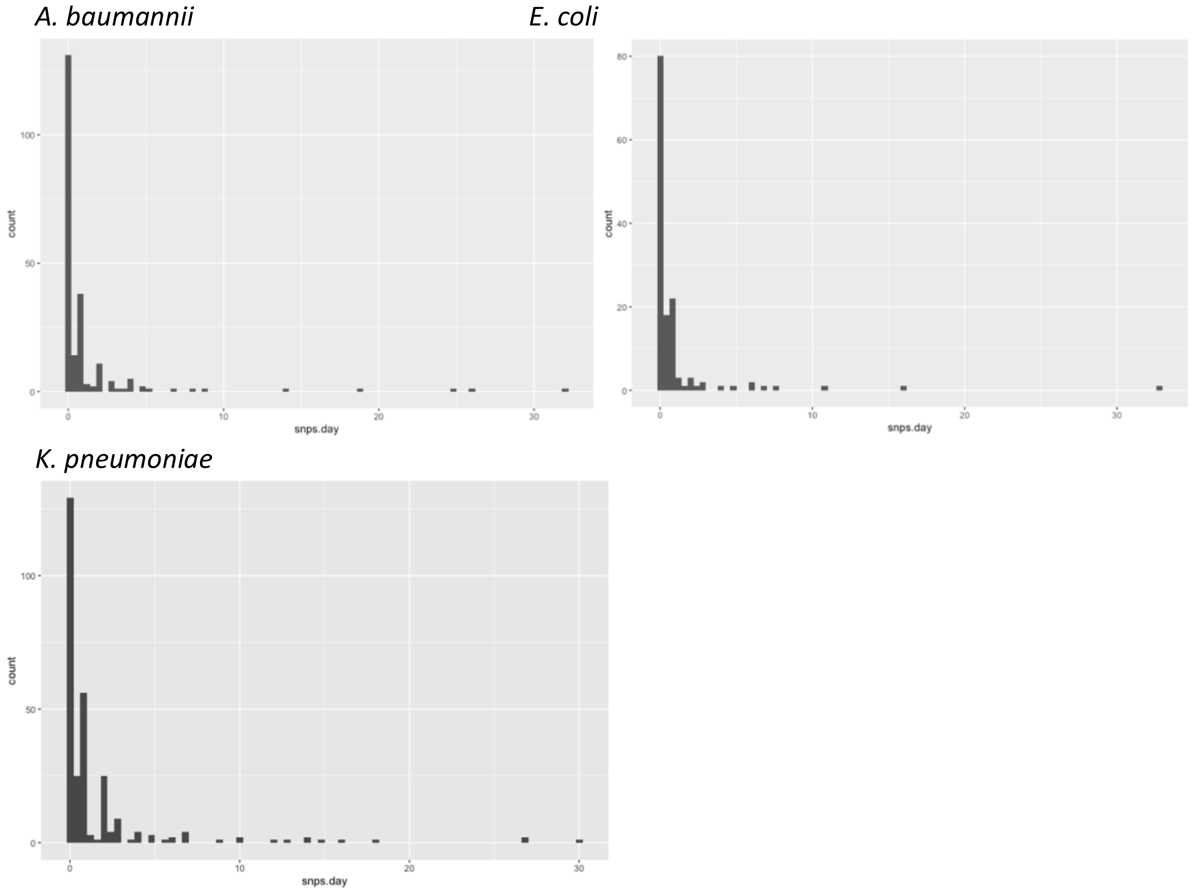


**Supplementary Figure 1: Summary of intra-patient basic SNP rates between isolates of the same species from the same patient.** Top left: *A. baumannii*, top right: *E. coli*, bottom: *K. pneumoniae*. The basic SNP rate is calculated by taking the first and last sample of the same species (and same ST) from a patient and calculating the number of SNPs divided by the number of days between sample isolation.

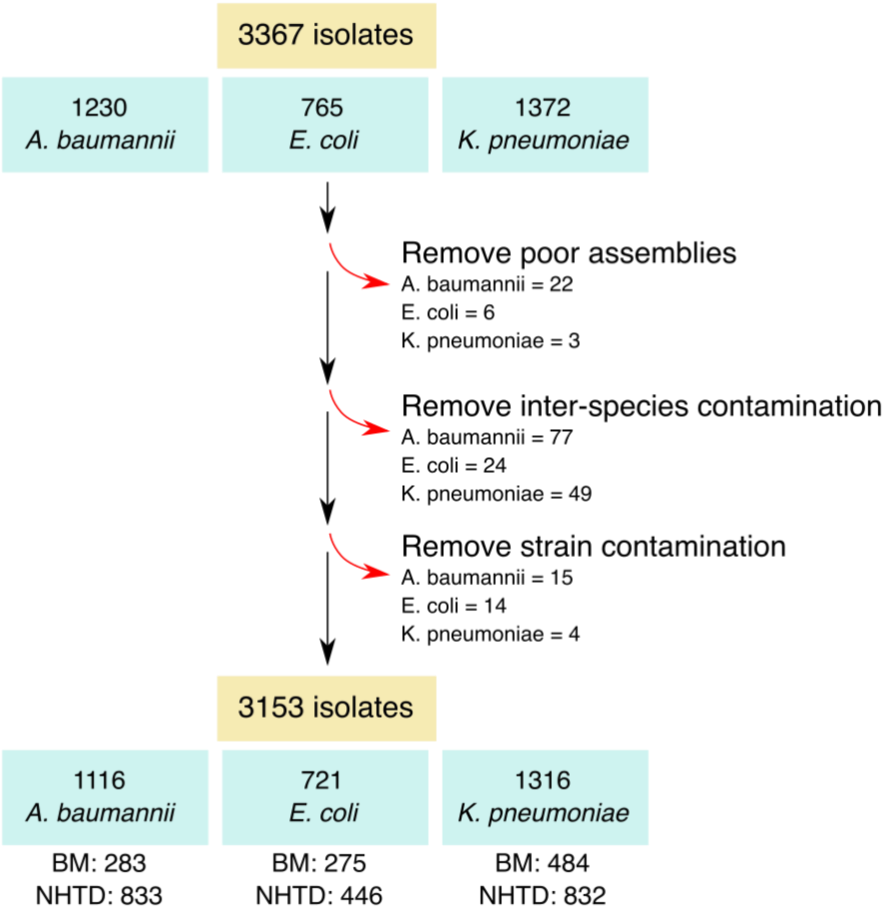


**Supplementary Figure 2: Isolate *de novo* assembly quality control process**

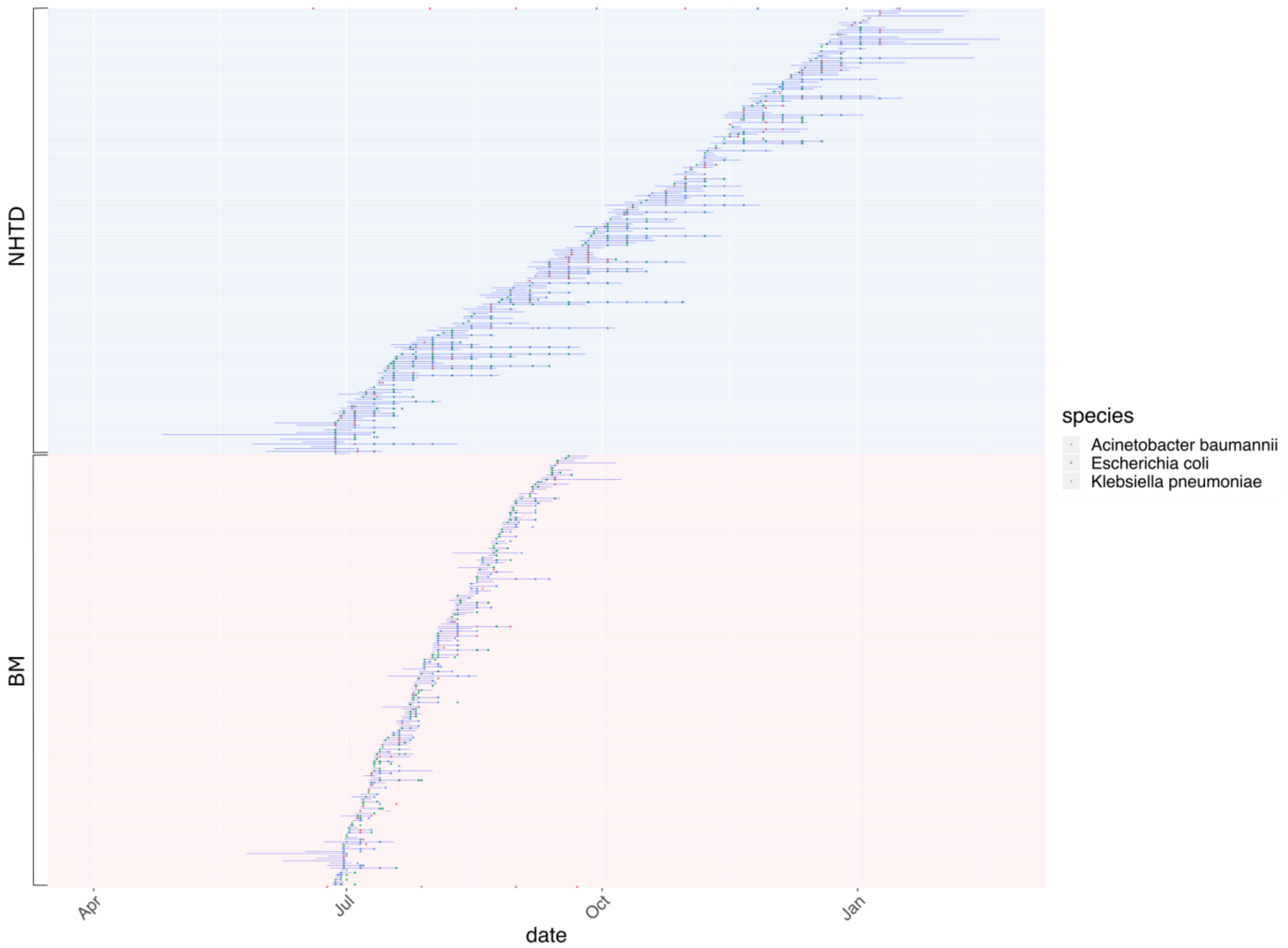


**Supplementary Figure 3: Timelines and isolate detection dates for all patients and samples in study (by hospital).** NHTD = National Hospital for Tropical Diseases. BM = Bach Mai Hospital. Solid blue lines represent admission ICU timeline for each patient.

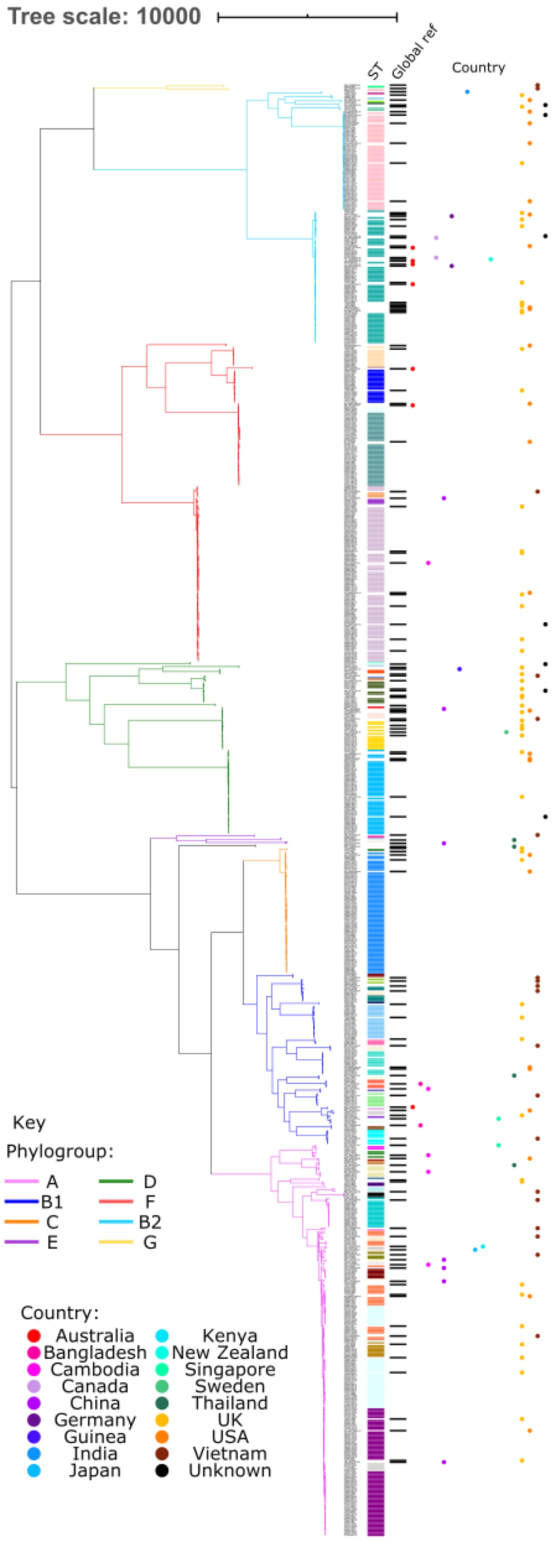


**Supplementary Figure 4: *E. coli* phylogeny showing study samples and global reference strains.** The columns to the right of the tree indicate ST (coloured bars), global reference strain (black bar) and country of origin (coloured dots)

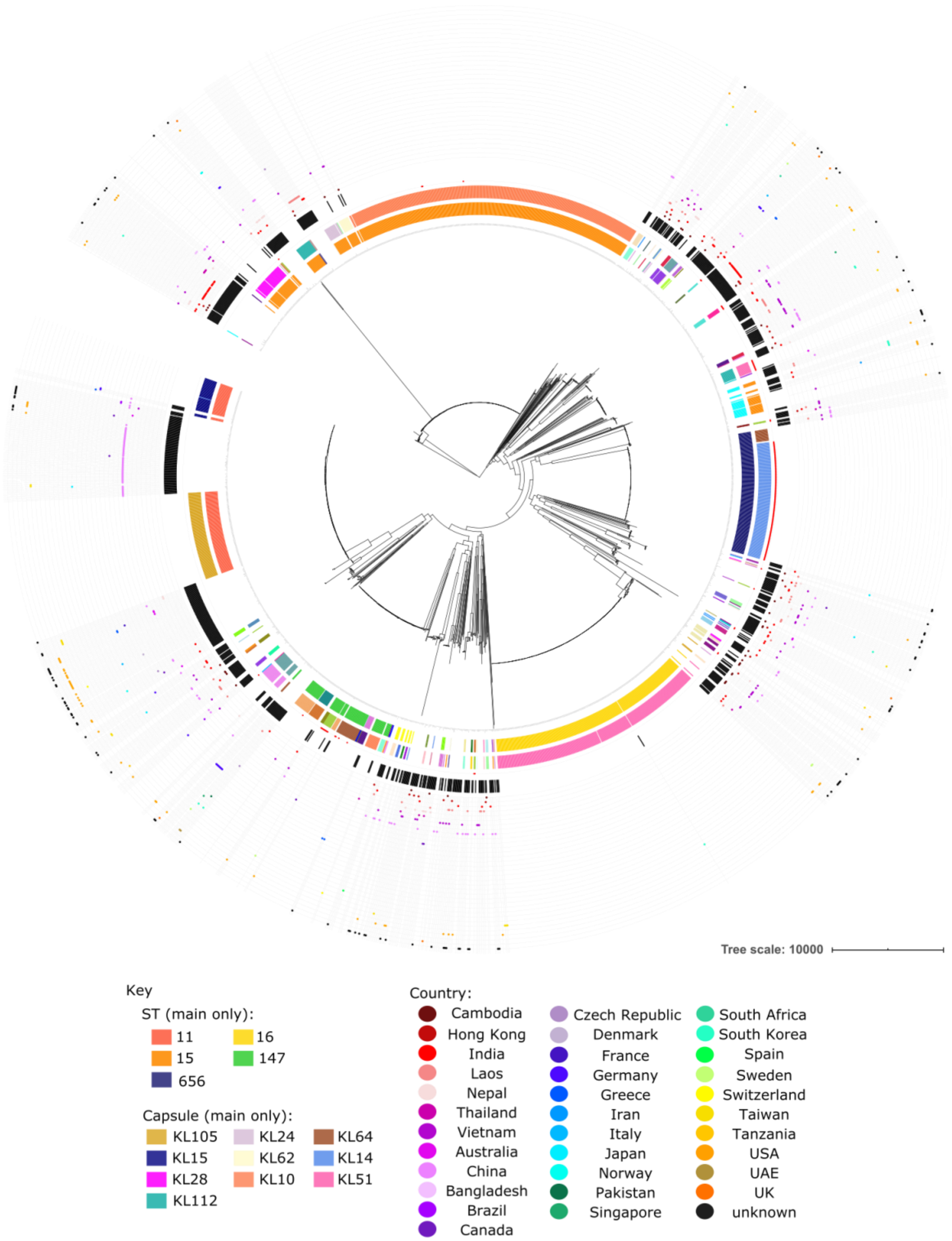


**Supplementary Figure 5:** ***K. pneumoniae*** **phylogeny of study isolates and global reference strains.** Rings annotated from innermost to outermost as follows: ring 1= multi-locus sequence type (MLST); ring 2 = capsule type; ring 3= capsule type confidence (red dot means low or no confidence, as per Kaptive result); ring 4 = global reference (black bars); outer rings = global reference countries (colored dots). Only the most prevalent capsule types/sequence types (STs) are shown.

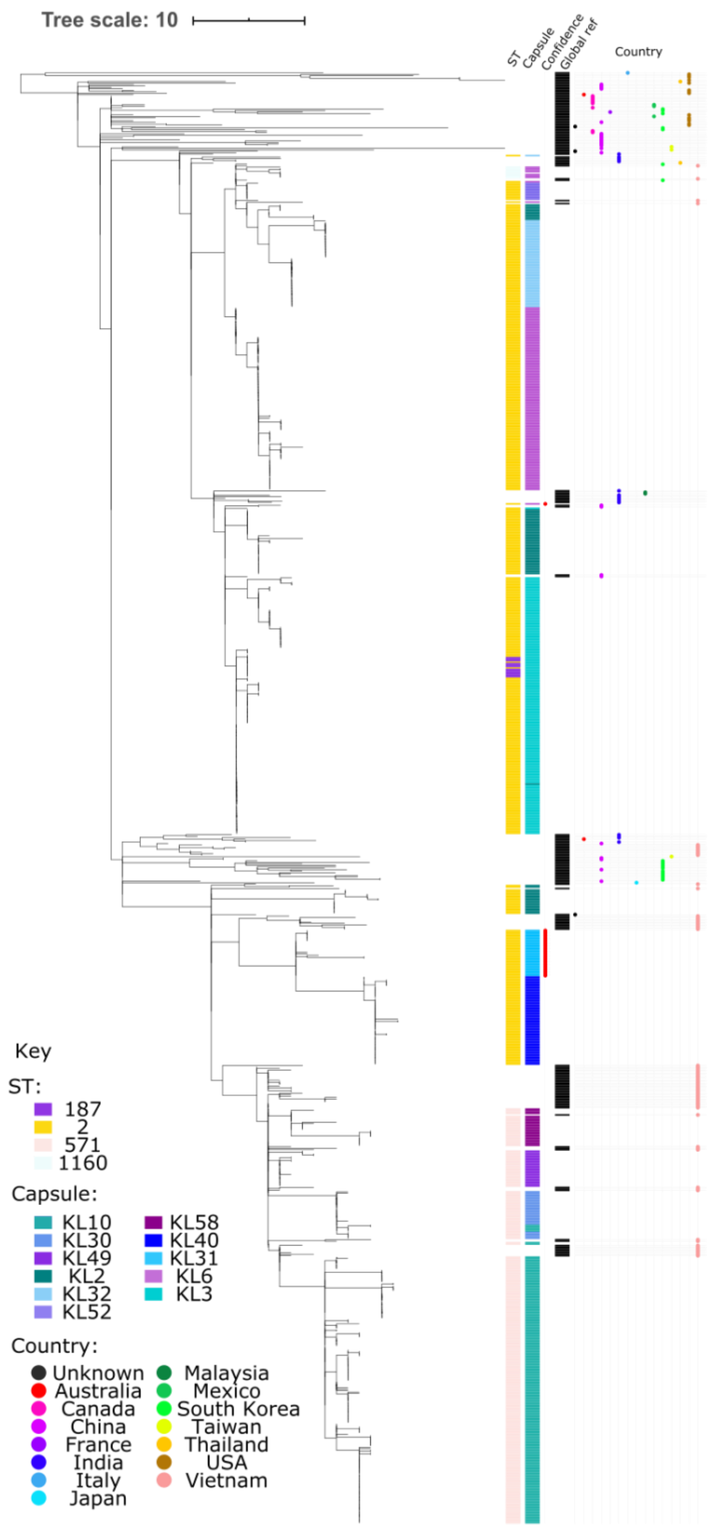


**Supplementary Figure 6: *A. baumannii* global clone 2 phylogeny with study isolates and global references.** The columns to the right of the tree indicate ST, capsule type, capsule type confidence (red dot = "low" or no confidence, as determined by Kaptive); global reference (black bar); country of origin (coloured dot).

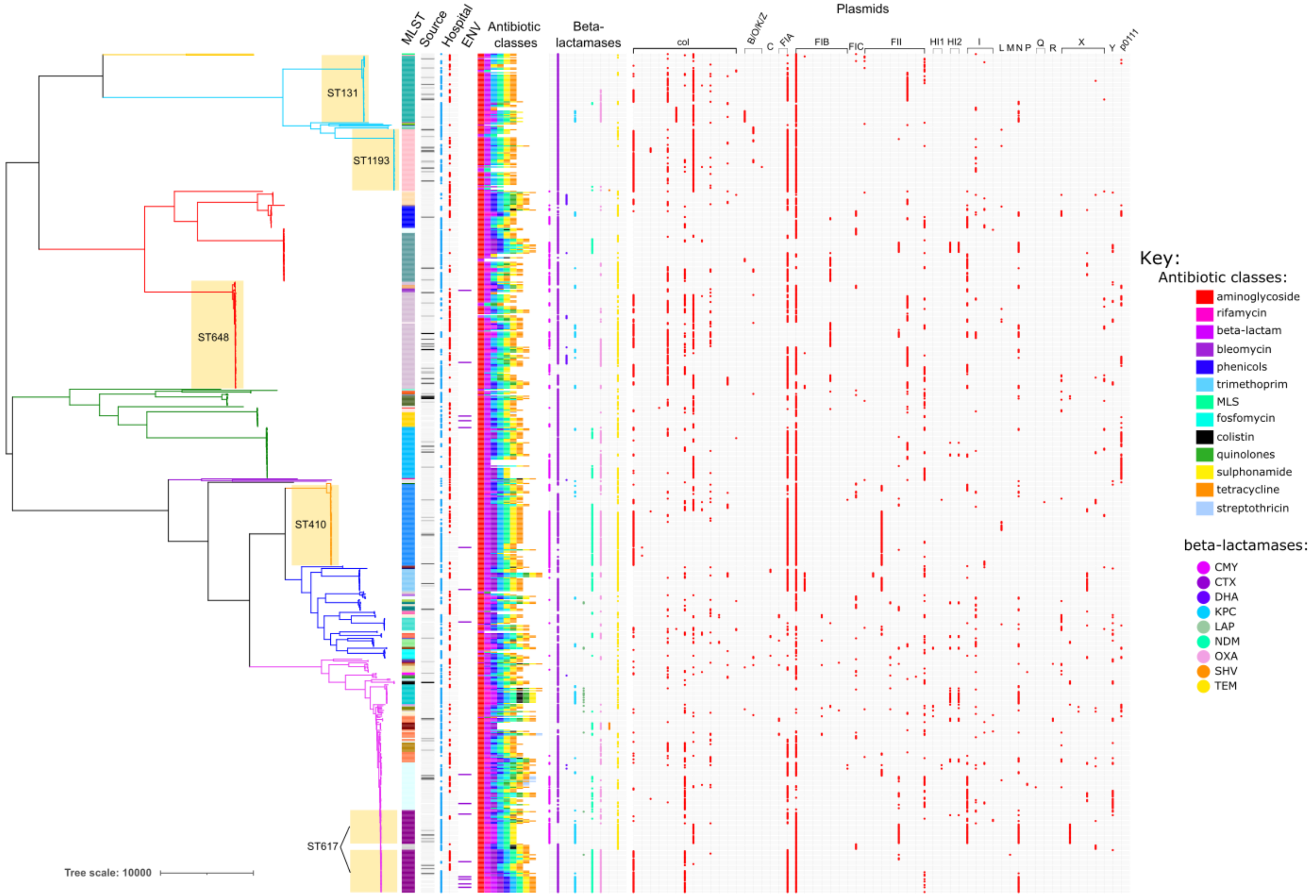


**Supplementary Figure 7: *E. coli* phylogeny annotated with acquired antibiotic resistance classes and plasmid incompatibility types.** Metadata columns annotated as follows: multi-locus sequence type (MLST); isolate source (as per Figure 1 main text); hospital (red =BMH, blue = NHTD); ENV environmental sample (purple bar); acquired resistance to antibiotic classes; beta-lactamase presence/absence; plasmid Inc types (red = present). MLS = macrolide-lincosamide-streptogramin. Main STs annotated on tree in yellow box.

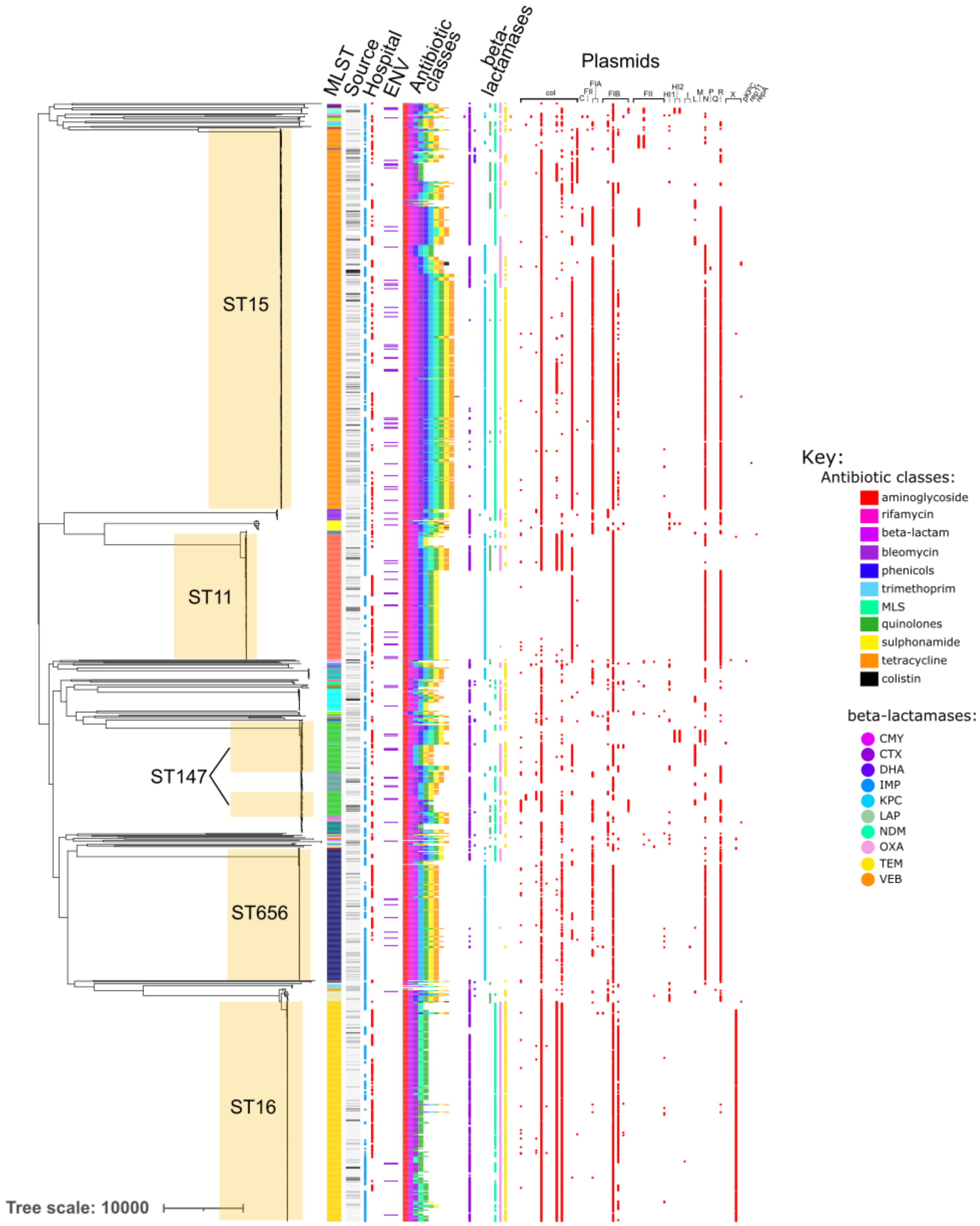


**Supplementary Figure 8: *K. pneumoniae* phylogeny annotated with acquired antibiotic resistance classes and plasmid incompatibility types.** Metadata columns annotated as follows: multi-locus sequence type (MLST); isolate source (as per Figure 1 main text); hospital (red =BMH, blue = NHTD); ENV environmental sample (purple bar); acquired resistance to antibiotic classes; beta-lactamase presence/absence; plasmid Inc types (red = present). MLS = macrolide-lincosamide-streptogramin. Main STs annotated on tree in yellow box.

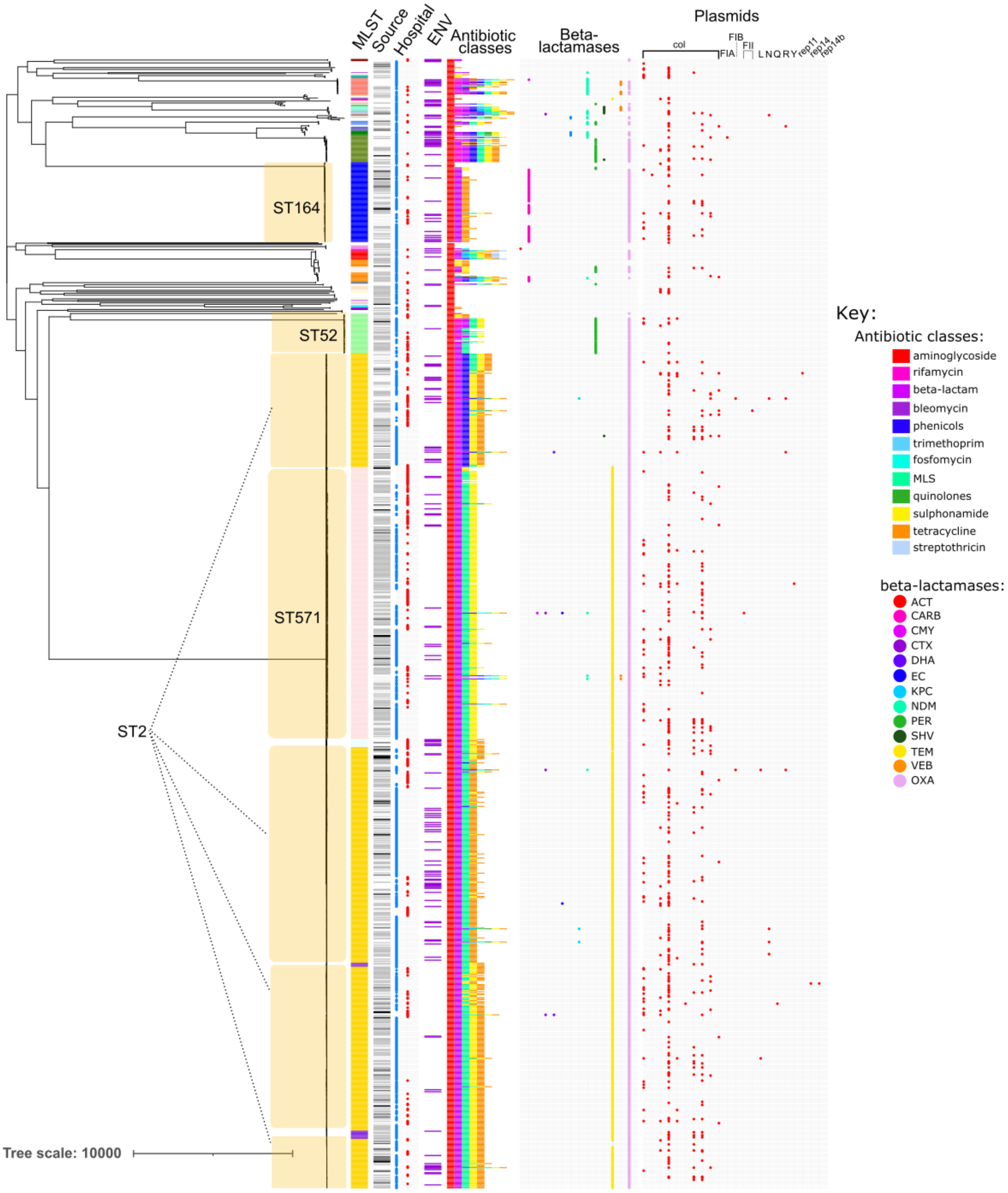


**Supplementary Figure 9: *A. baumannii* phylogeny annotated with acquired antibiotic resistance classes and plasmid incompatibility types.** Metadata columns annotated as follows: multi-locus sequence type (MLST); isolate source (as per Figure 1 main text); hospital (red =BMH, blue = NHTD); ENV environmental sample (purple bar); acquired resistance to antibiotic classes; beta-lactamase presence/absence; plasmid Inc types (red = present). MLS = macrolide-lincosamide-streptogramin. Main STs annotated on tree in yellow box.

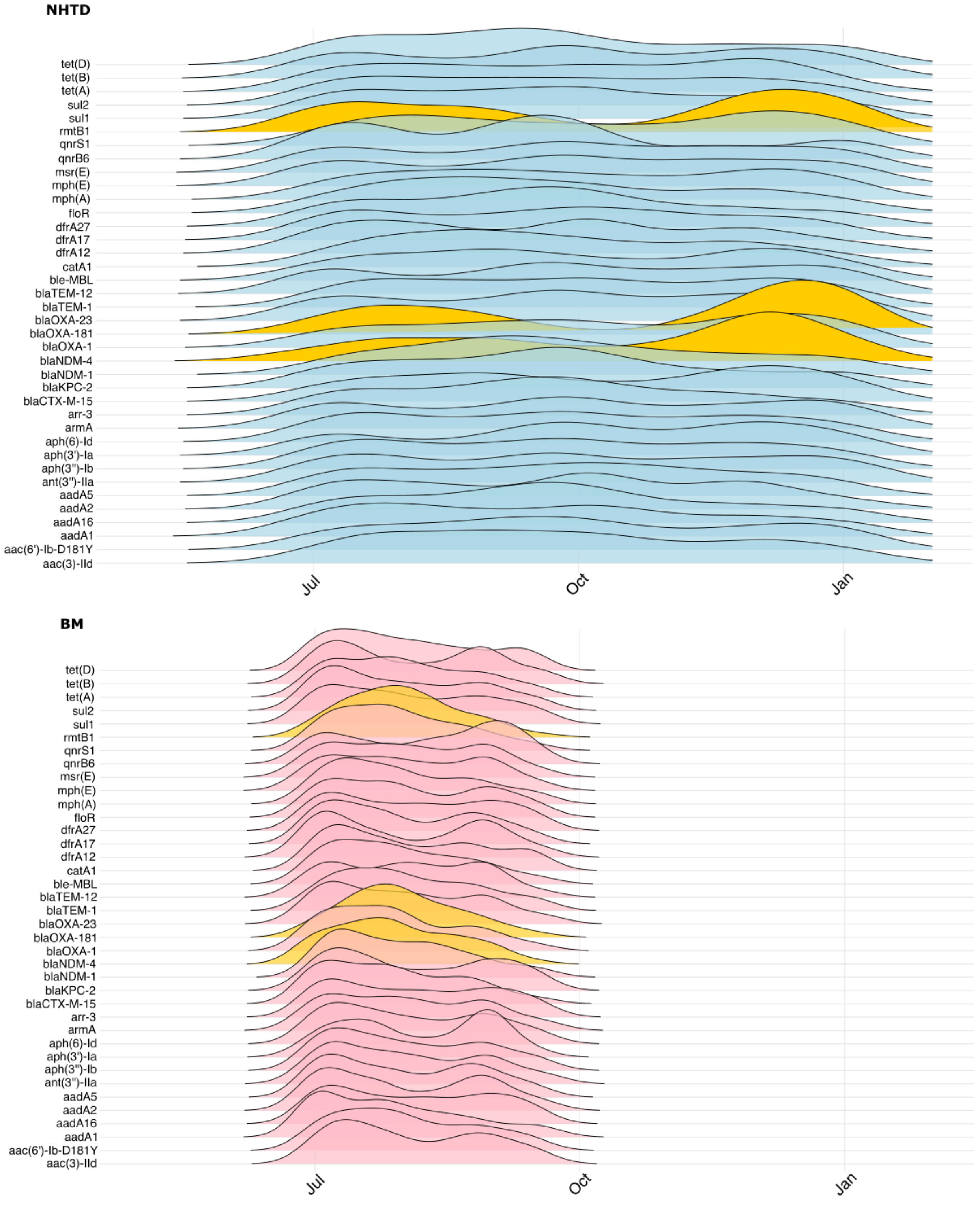


**Supplementary Figure 10: Gene presence versus date for all genes with ≥1% prevalence overall in either hospital.** NHTD samples (blue) in the upper panel and BMH samples (red) in the lower panel. Peaks coloured in orange (*rmtB1*, *bla*OXA-181, *bla*NDM-4) indicate genes that were found to have a similar increase in prevalence at a specific timepoint during the study period.

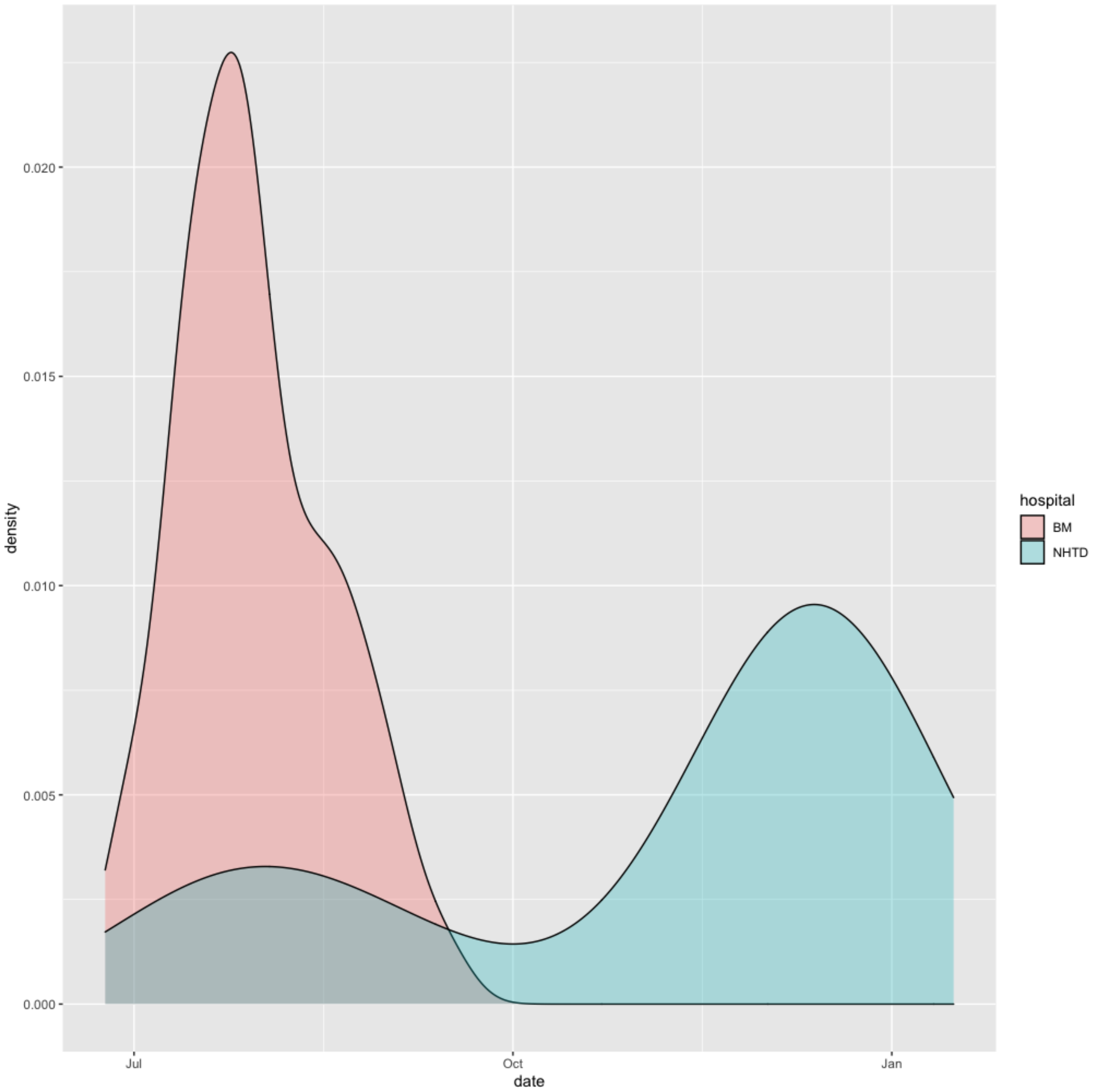


**Supplementary Figure 11: Date of isolation for *K. pneumoniae* isolates carrying *bla*NDM-4, *bla*OXA-181 and *rmtB1*.** BMH isolates are indicated in red and NHTD isolates in blue.

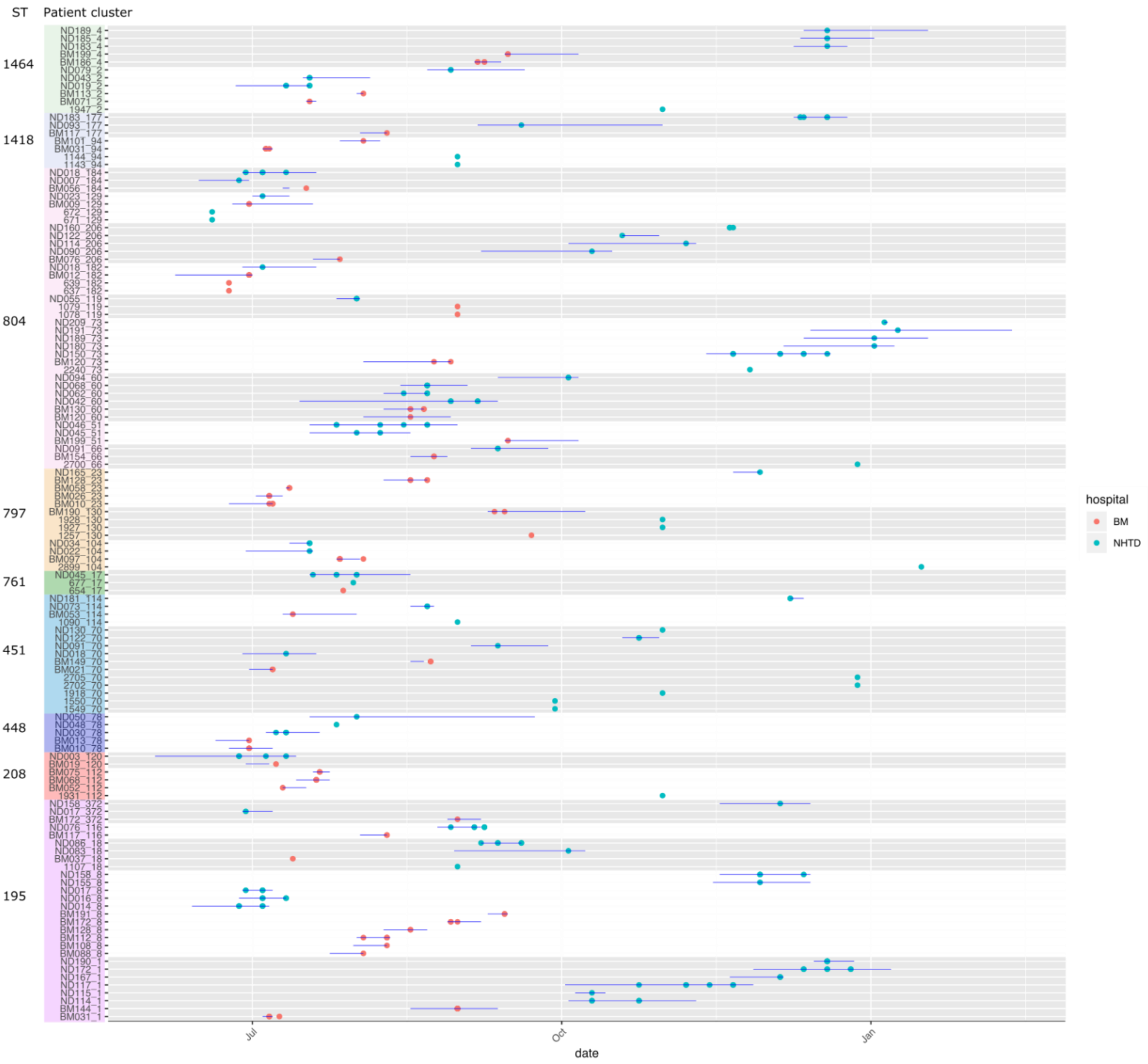


**Supplementary Figure 12: Epidemiological timelines for ICU patients positive for *Acinetobacter baumannii*, showing isolates involved in 0 SNP transmission clusters across both hospitals.** Each row is a patient, ordered by cluster (patients and cluster number described on y-axis as [PATIENT]_[CLUSTER_NUMBER], clusters are coloured by sequence type (Oxford scheme), as indicated to the left of the y axis). Alternating grey/white background separate clusters. Horizontal blue lines indicate individual patient ICU admission spells. Dots represent date of isolation of the patient strain involved in that cluster (coloured by hospital, red =BMH, blue =NHTD). Environmental isolates are labelled as numbers on the y-axis (no horizontal blue line).

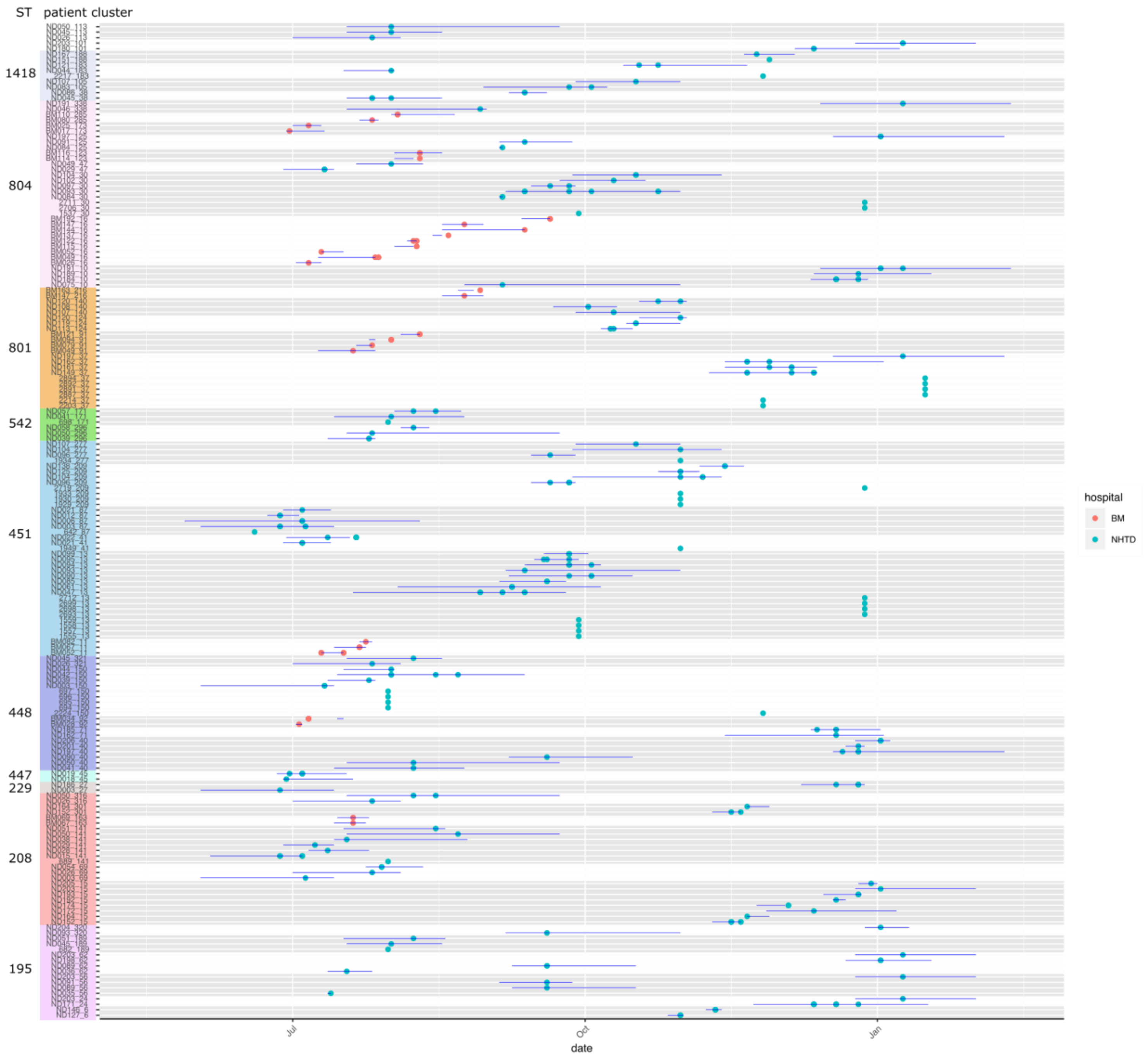


**Supplementary Figure 13: Epidemiological timelines for ICU patients positive for *Acinetobacter baumannii*, showing isolates involved in 0 SNP transmission clusters from a single hospital.** Each row is a patient, ordered by cluster (patients and cluster number described on y-axis as [PATIENT]_[CLUSTER_NUMBER], clusters are coloured by sequence type (Oxford scheme), as indicated to the left of the y axis). Alternating grey/white background separate clusters. Horizontal blue lines indicate individual patient ICU admission spells. Dots represent date of isolation of the patient strain involved in that cluster (coloured by hospital, red = BMH, blue = NHTD). Environmental isolates are labelled as numbers on the y-axis (no horizontal blue line).

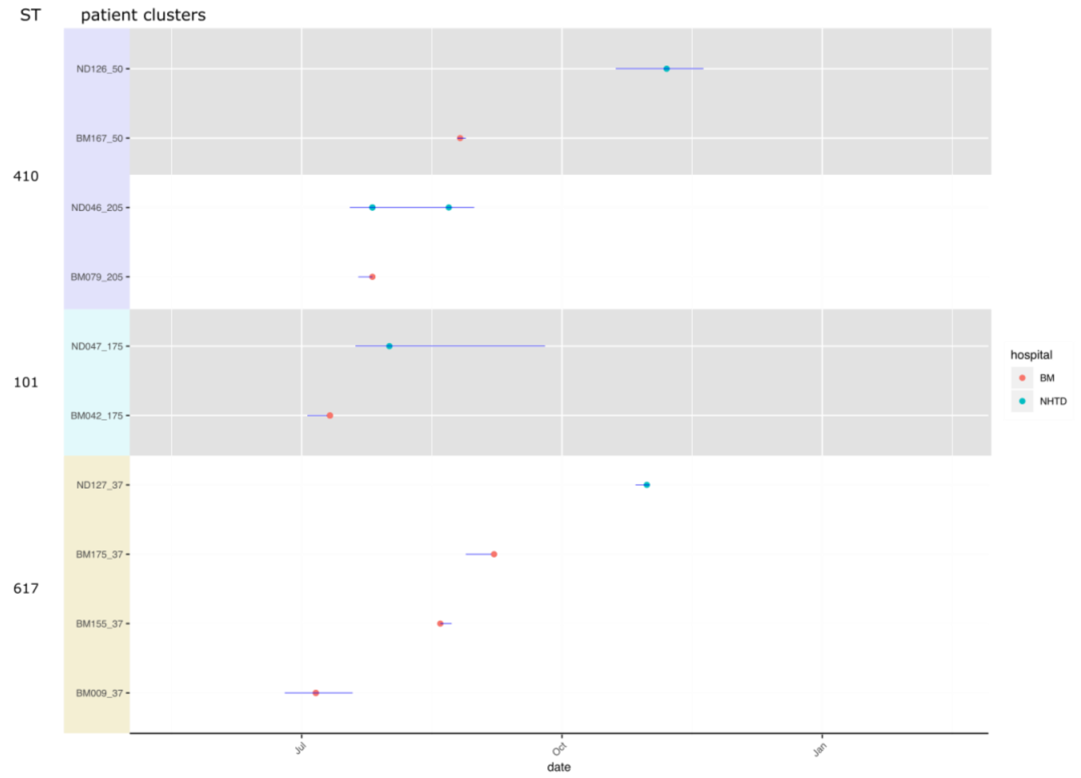


**Supplementary Figure 14: Epidemiological timeline of ICU patients positive for *Escherichia coli*, showing isolates involved in 0 SNP transmission clusters across both hospitals.** Each row is a patient, ordered by cluster (patients and cluster number described on y-axis as [PATIENT]_[CLUSTER_NUMBER], clusters are coloured by sequence type, as indicated to the left of the y axis). Alternating grey/white background separate clusters. Horizontal blue lines indicate individual patient ICU admission spells. Dots represent date of isolation of the patient strain involved in that cluster (coloured by hospital, red = BMH, blue = NHTD). Environmental isolates are labelled as numbers on the y-axis (no horizontal blue line).

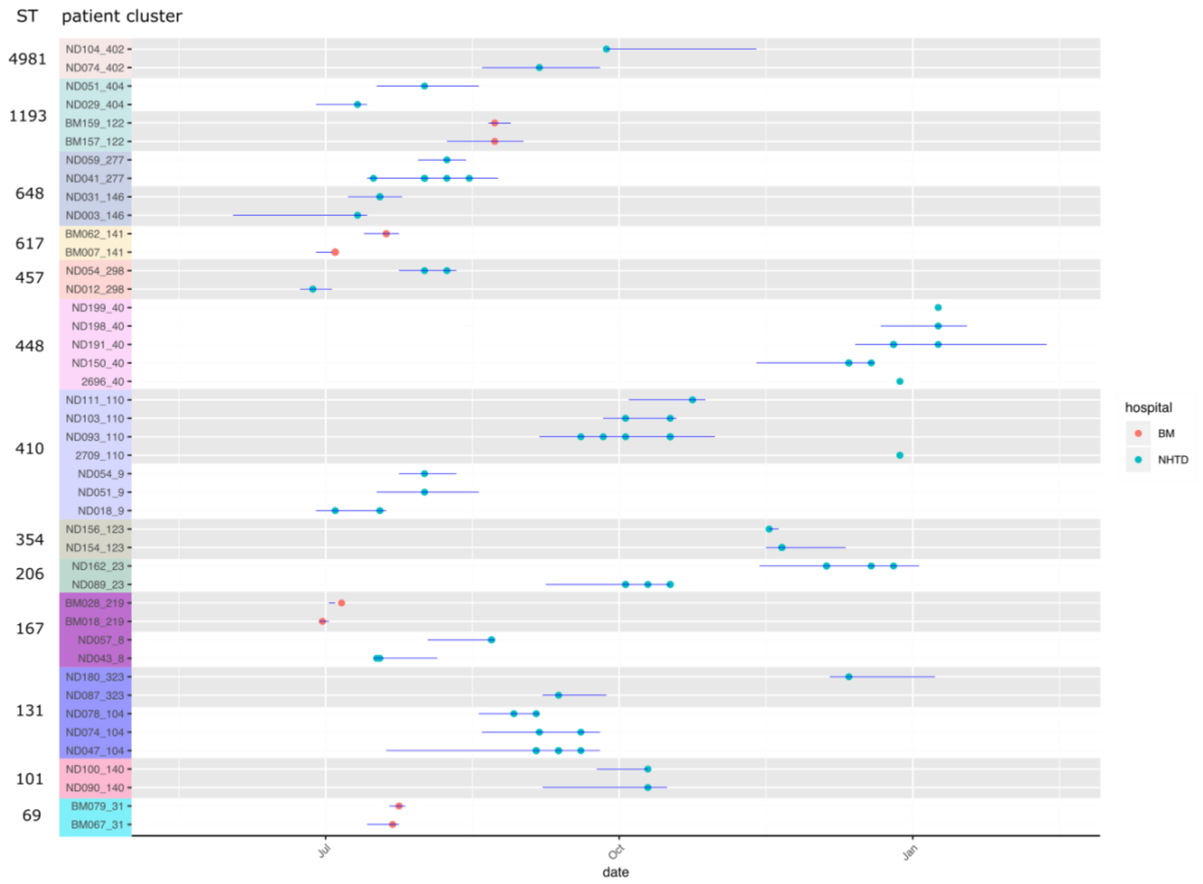


**Supplementary Figure 15: Epidemiological timeline of ICU patients positive for *Escherichia coli*, showing isolates involved in 0 SNP transmission clusters from a single hospital.** Each row is a patient, ordered by cluster (patients and cluster number described on y-axis as [PATIENT]_[CLUSTER_NUMBER], clusters are coloured by sequence type (Oxford scheme), as indicated to the left of the y axis). Alternating grey/white background separate clusters. Horizontal blue lines indicate individual patient ICU admission spells. Dots represent date of isolation of the patient strain involved in that cluster (coloured by hospital, red = BMH, blue = NHTD). Environmental isolates are labelled as numbers on the y-axis (no horizontal blue line).

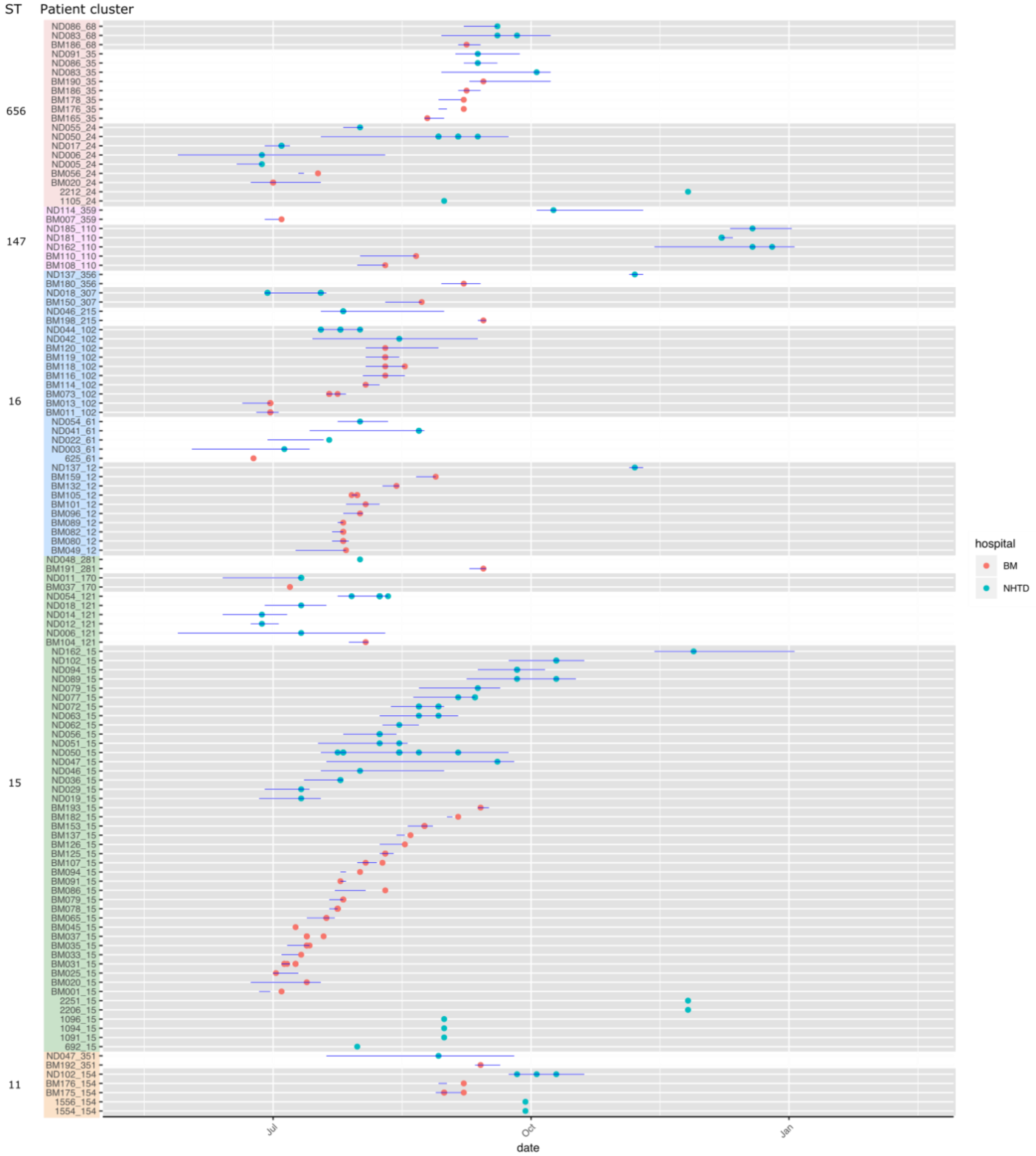


**Supplementary Figure 16: Epidemiological timeline of ICU patients positive for *Klebsiella pneumoniae*, showing isolates involved in 0 SNP transmission clusters across both hospitals.** Each row is a patient, ordered by cluster (patients and cluster number described on y-axis as [PATIENT]_[CLUSTER_NUMBER], clusters are coloured by sequence type, as indicated to the left of the y axis). Alternating grey/white background separate clusters. Horizontal blue lines indicate individual patient ICU admission spells. Dots represent date of isolation of the patient strain involved in that cluster (coloured by hospital). Environmental isolates are labelled as numbers on the y-axis (no horizontal blue line).

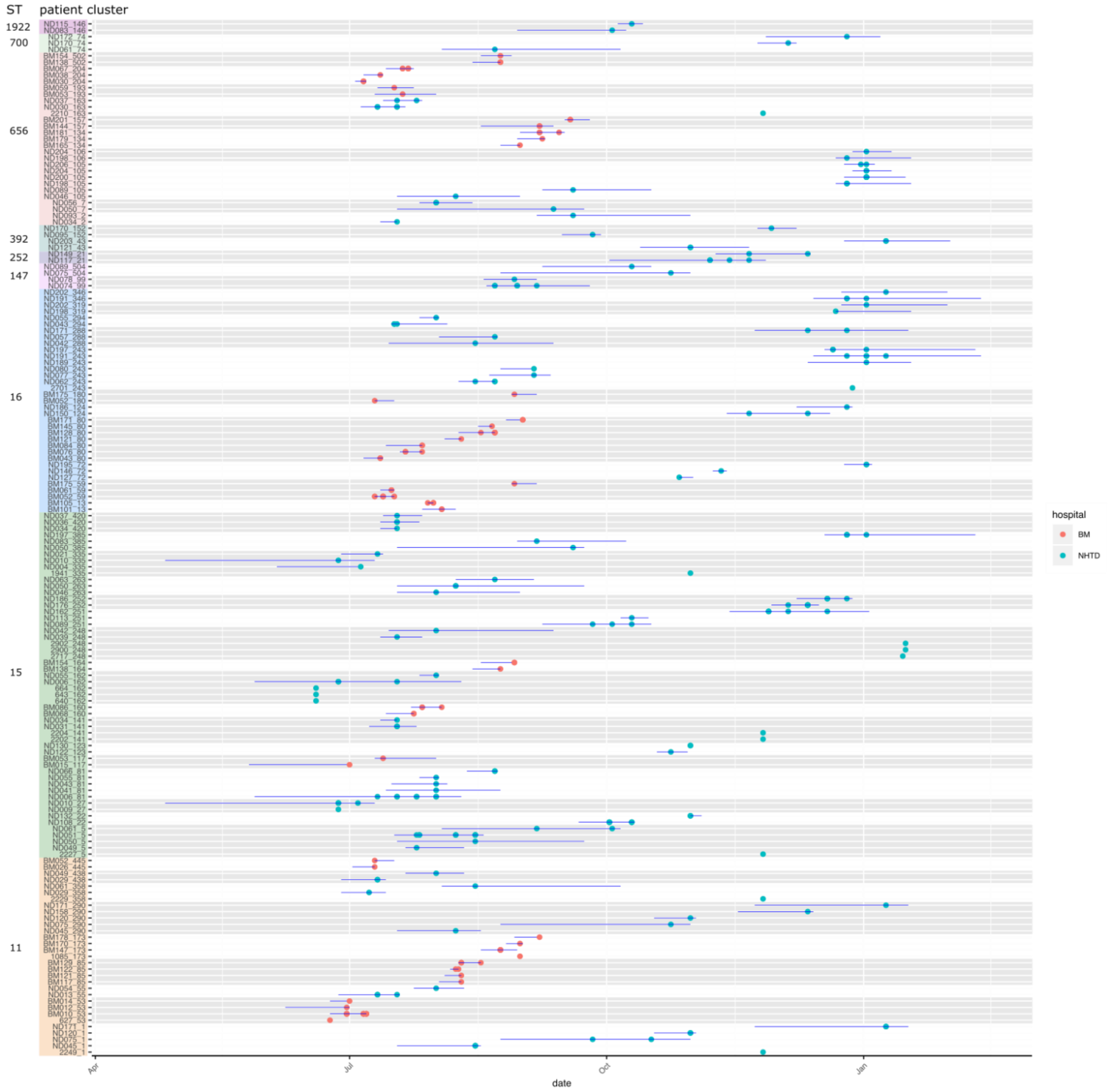


**Supplementary Figure 17: Epidemiological timeline of ICU patients positive for *Klebsiella pneumoniae*, showing isolates involved in 0 SNP transmission clusters from a single hospital.** Each row is a patient, ordered by cluster (patients and cluster number described on y-axis as [PATIENT]_[CLUSTER_NUMBER], clusters are coloured by sequence type (Oxford scheme), as described to the left). Alternating grey/white background separate clusters. Horizontal blue lines indicate individual patient ICU admission spells. Dots represent date of isolation of the patient strain involved in that cluster (coloured by hospital, red = BMH, blue = NHTD). Environmental isolates are labelled as numbers on the y-axis (no horizontal blue line).

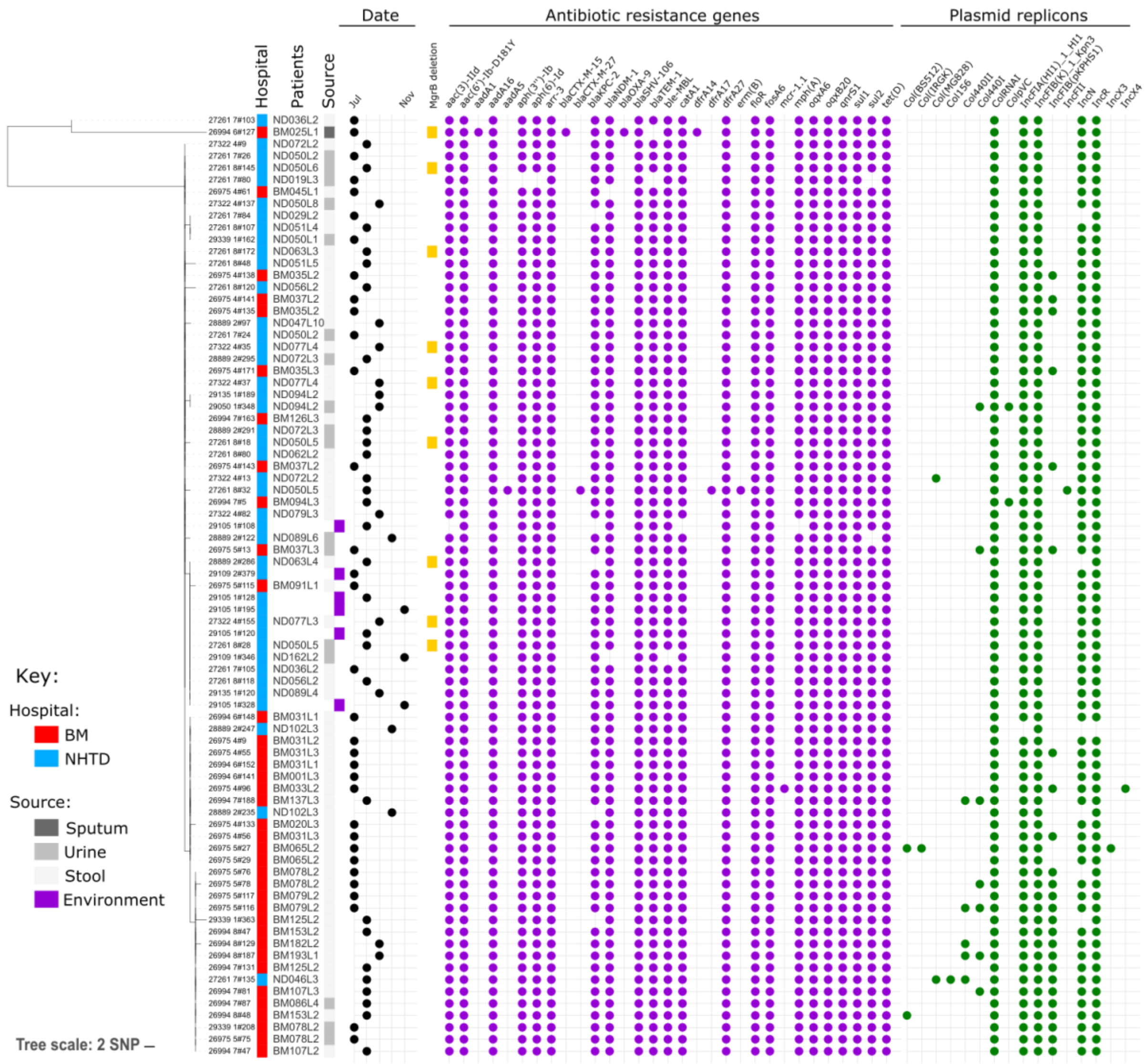


**Supplementary Figure 18: *Klebsiella pneumoniae* ST15 cluster.** The largest predicted transmission cluster identified was an ST15 cluster involving both ICU wards and 38 patients. Separate analysis of this cluster found that the majority of isolates differed by no more than 2 core SNPs, with the exception of two isolates that had a number of SNPs at a single gene locus (data not shown). All isolates shared similar plasmid replicons and antibiotic resistance gene profiles.

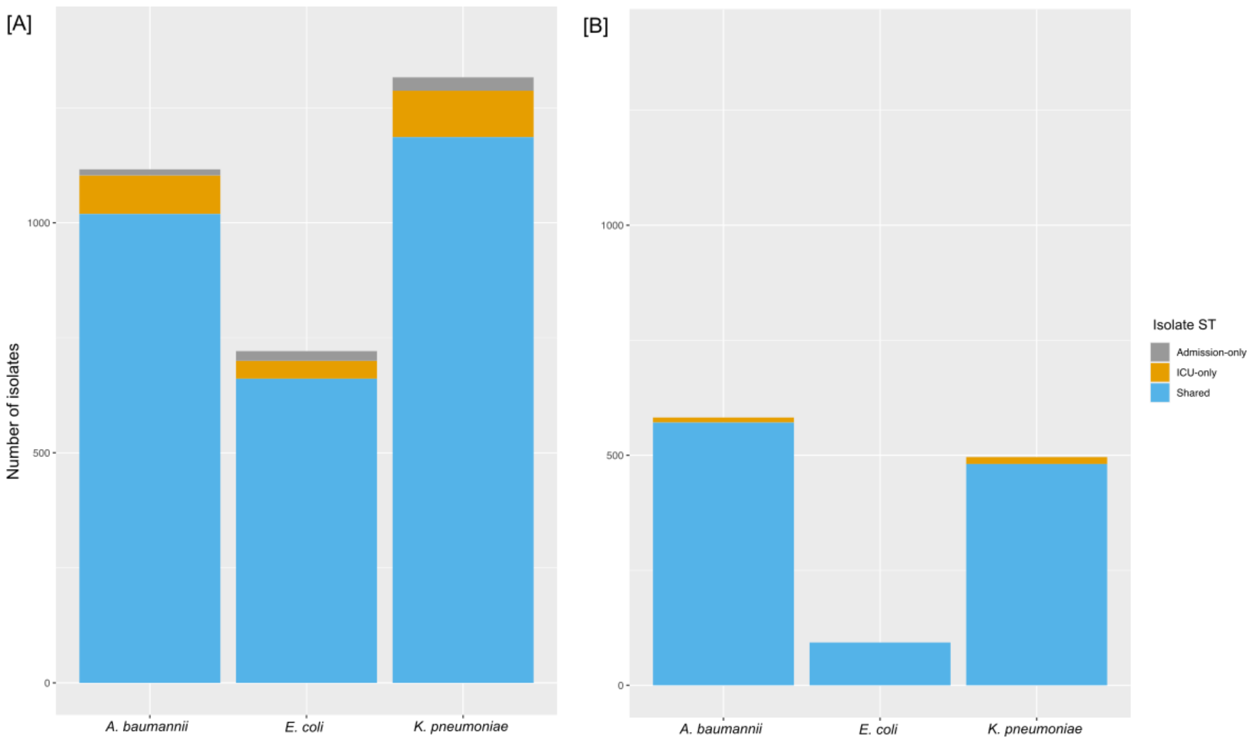


**Supplementary Figure 19: Breakdown of isolates belonging to sequence types (STs)** (i) only found on admission, (ii) only found in the ICU, or (iii) across both admission and ICU sampling points: We have used the following definitions, *Admission* = patient sample taken on admission to the ICU (first patient sample). *ICU* = patient sample taken once already residing in the ICU. [A] Count of all isolates across all three species coloured by STs that were isolated (i) only on admission (Grey), (ii) only within the ICU (orange) or (iii) shared - both on admission and within the ICU (blue). [B] Count of isolates involved in 0-SNP transmission clusters across all three species coloured by STs that were isolated (i) only within the ICU (orange) or ii) both on admission and within the ICU (blue). Notably, there were no transmission clusters from STs that were found only on admission.

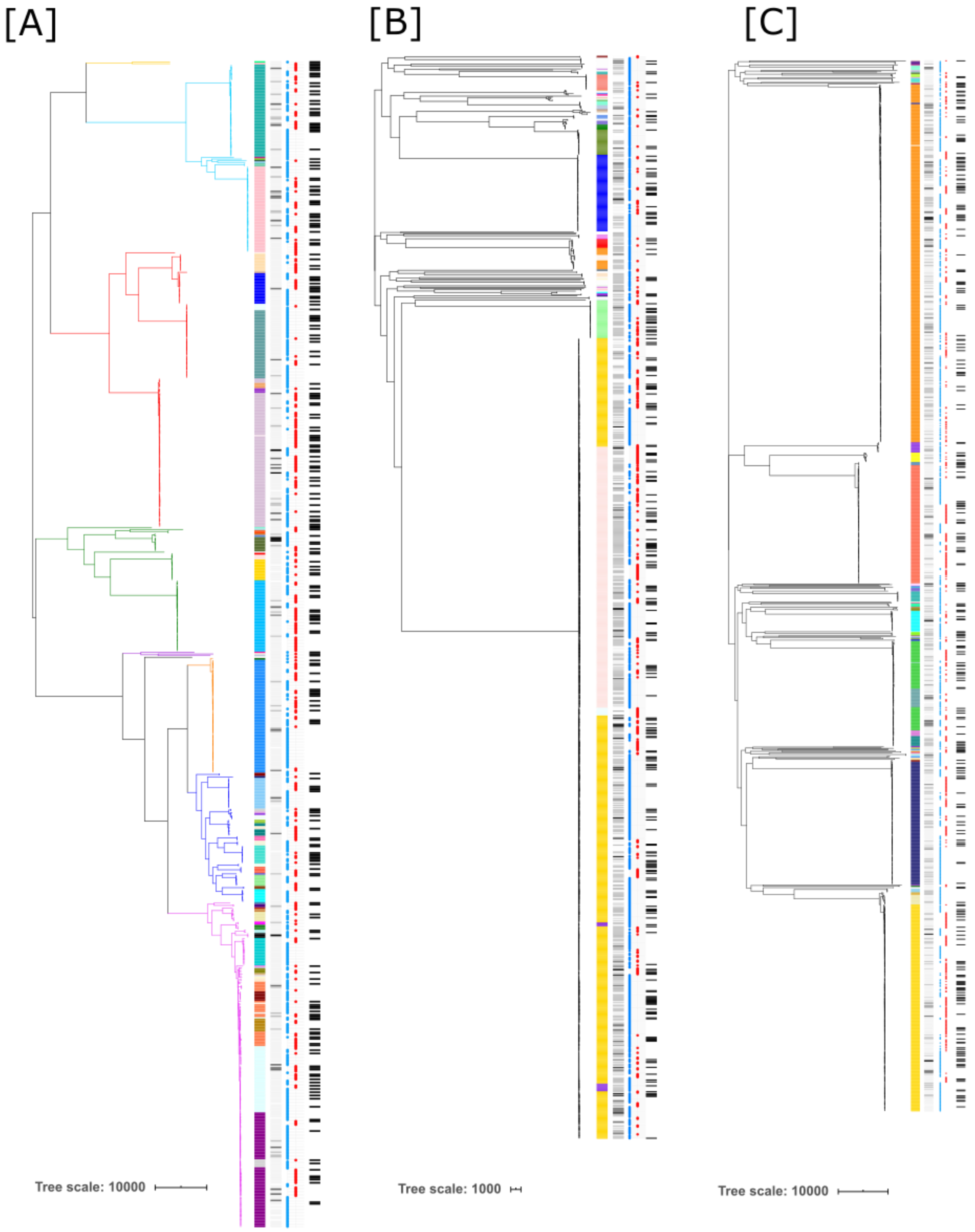


**Supplementary Figure 20:** Phylogenies of isolates detected on admission to ICU: [A] *E. coli*, [B] *A. baumannii*, [C] *K. pneumoniae*. Each tree is annotated with metadata columns from left to right: column 1= multi-locus sequence type (MLST); column 2 = sample type; column 3 = hospital (BMH = red, NHTD = blue); column 4 = isolate detected in admission sample (black bar). Admission samples were distributed throughout the phylogenies and no new clades emerged after ICU admission.

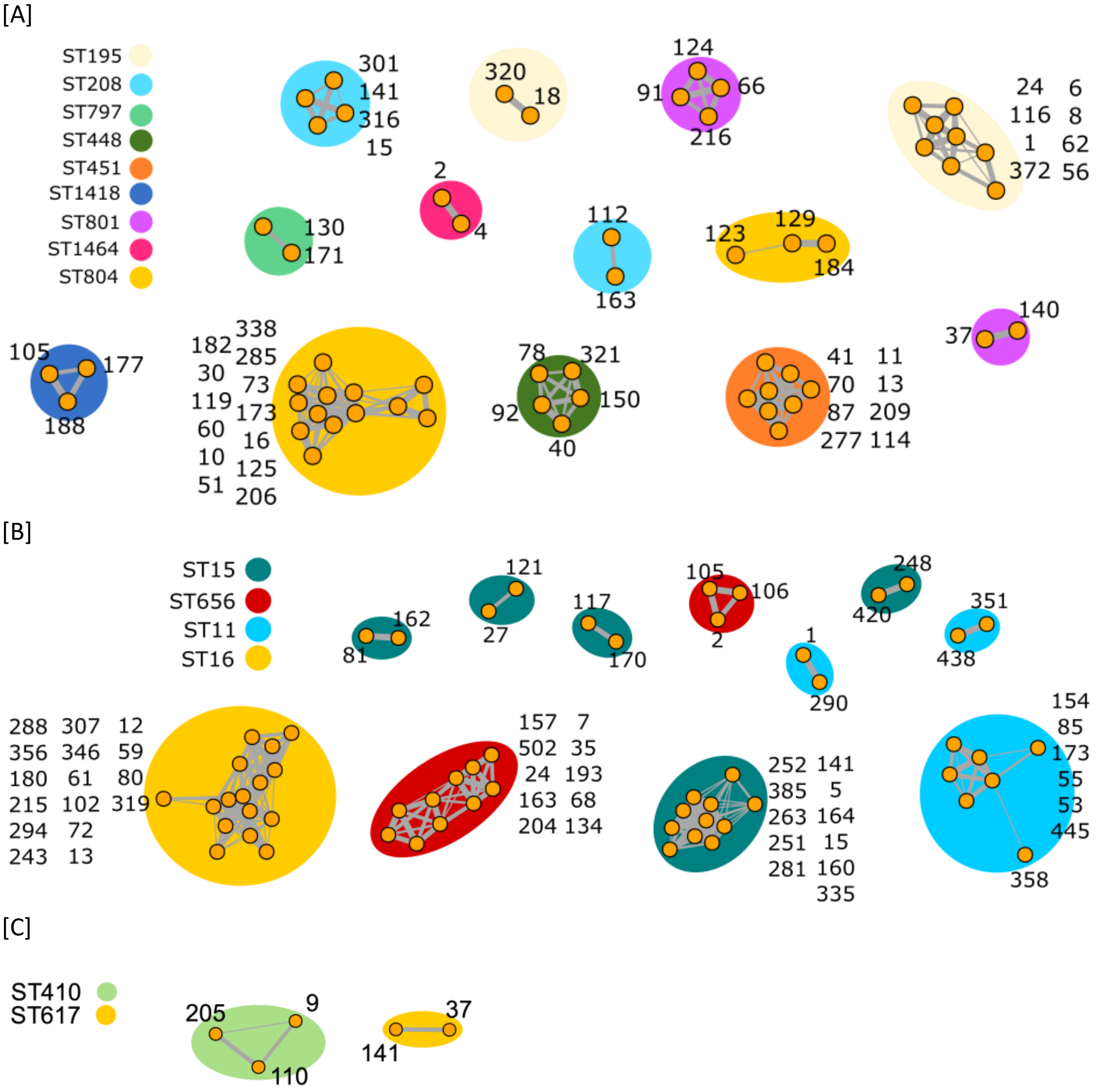


**Supplementary Figure 21:** Five SNP clustering of original (0 SNP) transmission clusters by organism [A] *A. baumannii*, [B] *K. pneumoniae*, [C] *E. coli*. Original 0 SNP clusters are represented by orange circles, where grey lines connecting them represent clustering at a 5 SNP threshold. The original assigned cluster numbers are written besides each circle. Background colours represent sequence types (STs).

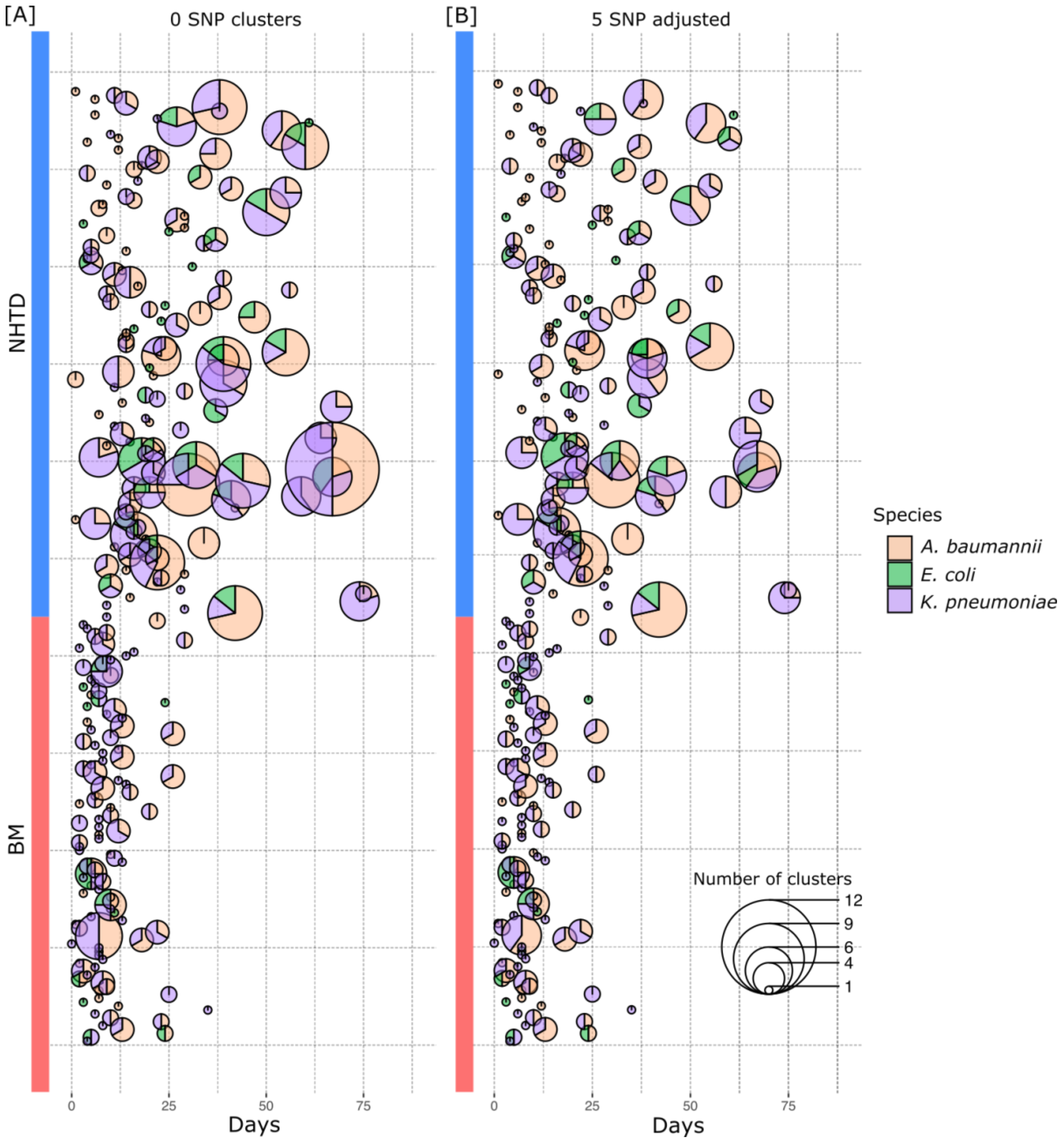


**Supplementary Figure 22: Scatterpie showing the number of clusters in patients across all species:** y-axis represents patients from BM or NHTD. X-axis represents length of stay for that patient; one pie is plotted per patient at the duration of their stay. **[A]** patients involved in at least one 0 SNP cluster. Each circle represents 0 SNP clusters involving a single patient. The size of each circle corresponds to the number of clusters involving that patient, while the colour corresponds to the species. **[B]** 5-SNP adjusted clusters. Each circle now shows how the original 0-SNP clusters merge at a 5-SNP threshold (i.e. clusters are now 5-SNP clusters). The size of the circle corresponds to the number of clusters, while the colour corresponds to the species.
